# Machine learning model predicts new-onset lower extremity deep vein thrombosis after pelvic fracture surgery and targeted diagnosis

**DOI:** 10.64898/2025.12.01.25341405

**Authors:** Haoyuan Fu, Qi Dong, Guoqiang LI, Kuo Zhao, Zhiyong Hou

**Author notes:** Correspondence: Dr. Zhiyong Hou, Department of Orthopedic Surgery, Third Hospital of Hebei Medical University, Shijiazhuang 050051, China., Dr. Kuo Zhao, Department of Orthopedic Surgery, Third Hospital of Hebei Medical University, Shijiazhuang 050051, China. Haoyuan Fu, Qi Dong, Guoqiang Li made equal contributions.

## Abstract

**Background:** Postoperative new-onset deep vein thrombosis (PNO-DVT) is a common and serious complication after pelvic fracture surgery, significantly affecting recovery and quality of life. Traditional risk assessment tools lack precision, whereas machine learning offers improved predictive capability. This study evaluated machine learning models for predicting PNO-DVT following pelvic fracture surgery.

**Methods:** Clinical data from 745 patients treated between 2016 and 2019 were analyzed. Demographic characteristics, laboratory indicators, surgical variables, and scoring systems were collected. Univariate logistic regression, LASSO regression, and multivariate logistic regression identified 12 independent risk factors. The dataset was split 7:3 into training and test sets. Six machine learning models (logistic regression, SVM, random forest, XGBoost, LightGBM, and AdaBoost, were constructed.

**Results:** Univariate analysis identified GGT, HDL-C, ApoB, TCOLJ, GLU, GAP, EOS, and MPV as potential predictors. Multivariate analysis confirmed age, nephropathy, BMI, fracture reduction, intraoperative blood loss, red blood cell transfusion, HDL-C, ApoB, GAP, and MPV as independent risk factors. Among all models, XGBoost achieved the highest AUC (0.8633), demonstrating superior predictive performance. Model accuracy ranged from 0.6502 to 0.9283.

**Conclusion:** Machine learning (particularly XGBoost) provides effective prediction of PNO-DVT after pelvic fracture surgery. Key predictors such as age, intraoperative blood loss, and BMI enable accurate risk stratification and support early preventive intervention.

## Background

Pelvic fractures represent a severe form of trauma associated with a broad spectrum of complex and diverse postoperative complications. Notably, the development of deep vein thrombosis (DVT) in the lower extremities is a critical factor contributing to elevated mortality and disability rates among these patients. Over the years, numerous studies have demonstrated that individuals with pelvic fractures are predisposed to a hypercoagulable state, attributed to factors such as prolonged immobilization, tissue damage, and postoperative inflammatory responses, thereby elevating the risk of DVT ^[1–2]^. The incidence of lower limb DVT in patients with pelvic fractures has been reported to reach 20% or higher, with most cases occurring postoperatively, thereby significantly impacting patient rehabilitation outcomes and quality of life ^[3–4].^

Traditional risk assessment for DVT predominantly relies on clinical experience and isolated clinical indicators, such as D-dimer levels, patient age, and body mass index, in conjunction with straightforward assessment scales, including the Wells score, Autar score, and Padua score. However, these methodologies exhibit notable limitations in practical application, characterized by inadequate predictive accuracy and specificity, impairing the ability to achieve early and precise risk stratification in clinical settings. For instance, while the Autar and Padua scores demonstrate high sensitivity in intensive care unit (ICU) populations, their low specificity contributes to a significant rate of misdiagnosis, thereby complicating the implementation of tailored preventive strategies^[5–6].^ Furthermore, there is a dearth of specialized risk assessment instruments for patients with pelvic fractures, as traditional models are predominantly grounded in single disease contexts, failing to account for the intricate pathological conditions associated with pelvic fractures^[2]^.

With the rapid advancement of artificial intelligence and machine learning, risk prediction models leveraging big data have demonstrated significant potential in the medical domain. Machine learning is adept at processing extensive, multidimensional clinical datasets, enabling automatic learning and identification of complex, nonlinear relationships, thereby enhancing the accuracy and stability of risk assessments. Recent studies have developed machine learning models to predict postoperative DVT risks in patients undergoing brain trauma, tumor surgeries, and multiple injuries, achieving commendable predictive performance. For instance, models utilizing algorithms such as Gradient Boosting Machine (GBM) and LightGBM have reported area under the curve (AUC) values of 0.85 or higher in predicting DVT following brain trauma and brain tumor surgeries, markedly outperforming traditional statistical models^[7–9]^. Furthermore, integrating feature selection via Support Vector Machine (SVM) with reinforcement learning algorithms has achieved an accuracy of 95.9% for DVT risk prediction, underscoring the distinct advantages of machine learning in extracting critical risk factors and optimizing prediction models ^[10]^.

Previous studies on pelvic fracture patients have sought to develop risk prediction models based on clinical variables. These include multifactorial regression models that integrate factors such as emergency abdominal surgery, the Injury Severity Score (ISS), and indicators of renal and liver function. The resulting risk assessment nomogram has demonstrated high predictive accuracy, with an AUC of approximately 0.88, in both training and validation cohorts, thereby serving as an effective tool for preoperative risk stratification of DVT^[2]^. Nonetheless, the comprehensive application of machine learning techniques to predict the risk of new-onset lower limb DVT following pelvic fracture surgery remains nascent. Accordingly, there is a pressing need to systematically incorporate large-scale, multicenter datasets and integrate multimodal clinical information to develop a more precise, clinically valuable prediction model with substantial clinical application potential.

Accordingly, a combination of traditional logistic regression and machine learning algorithms was adopted in the present study to develop a predictive model for new-onset lower limb DVT after pelvic fracture surgery, thereby enhancing clinical decision-making.

## Methods

### Data collection and processing

The study collected data from 857 patients diagnosed with pelvic fractures at the Third Hospital of Hebei Medical University from January 2016 to December 2019. DVT was confirmed by compression Doppler ultrasonography, primarily defined by venous non-compressibility alongside intraluminal echogenic material and absent flow variation. To minimize detection bias and accurately determine the temporal sequence of events, a standardized screening protocol was implemented for all eligible patients. This protocol included assessments at specific preoperative and postoperative intervals (days 3, 7, and 14). Consequently, blood indicators were tested both before and after the potential onset of thrombosis, allowing for a longitudinal analysis of biomarker changes and the inclusion of both asymptomatic and symptomatic cases to accurately reflect the true incidence within the cohort. Given that the samples were collected across multiple time intervals, we were able to distinguish between characteristics that existed before thrombosis and those that emerged after thrombosis. For characteristics tested after thrombosis, we considered that they might reflect the pathophysiological state of the disease rather than purely predictive functions. This analysis specifically evaluated whether the selected characteristics, especially those measured before thrombosis, possess the ability to serve as early predictive factors, rather than merely correlating with the established disease. After excluding 112 patients with preoperative thrombosis, 745 patients who developed postoperative DVT were included in the final analysis. The incidence of postoperative non-occlusive deep vein thrombosis (PNO-DVT) was 8.5% (n=64/681), with a predominance of male patients (74.8%, n=558/857).

### Identification of risk factors for PNO-DVT

Independent risk factors were identified using a two-step analytical approach. Initially, a single-factor logistic regression analysis was employed to select factors with a *P*-value ≤ 0.05. Subsequently, the least absolute shrinkage and selection operator (LASSO) regression was utilized to further refine the selection of influencing factors for postoperative thrombosis, addressing potential collinearity issues. In the LASSO regression model, an increasing λ value intensified the penalty term, driving the coefficients of the independent variables towards zero. This regularization process effectively mitigated model overfitting. Based on the nine variables identified through univariate analysis and LASSO regression, a multivariate logistic regression was conducted to identify independent predictors of postoperative thrombosis.

### Model establishment

The dataset for model establishment was derived from selecting 12 significant features identified through single-factor analysis. This dataset was then randomly partitioned into a training set (70%) and a test set (30%). Given the approximately 1:10 ratio of positive to negative samples, a notable class imbalance was present. To mitigate potential model bias towards the negative samples, which could lead to a diminished recall rate, sampling techniques were applied exclusively to the training set, leaving the test set unaltered. To further ensure the robustness and stability of the performance evaluation, 10-fold cross-validation was performed on the training set. Six machine learning algorithms, including logistic regression, SVM, random forest, XGBoost, LightGBM, and AdaBoost, were employed to develop six distinct models. To optimize model performance, hyperparameter tuning was performed using grid search optimization on the training set to identify the most effective parameter configurations for each algorithm. All statistical analyses and data visualizations were executed using R version 4.5.1. The study utilized the “xgboost”, “e1071”, “randomForest”, “rpart”, and “caret” packages for machine learning modeling, and the “tidyverse”, “pROC”, “ggplot2”, “gridExtra”, “tibble”, and “shapviz” packages for data organization and visualization.

## Results

### 1. Results of univariate logistic regression analysis

According to the univariate analysis of baseline characteristics in Table 1, age, hypertension, kidney disease, rheumatoid disease, peripheral vascular disease, and body mass index (BMI) were identified as potential risk factors for the development of postoperative thrombosis.

**Table 1:**
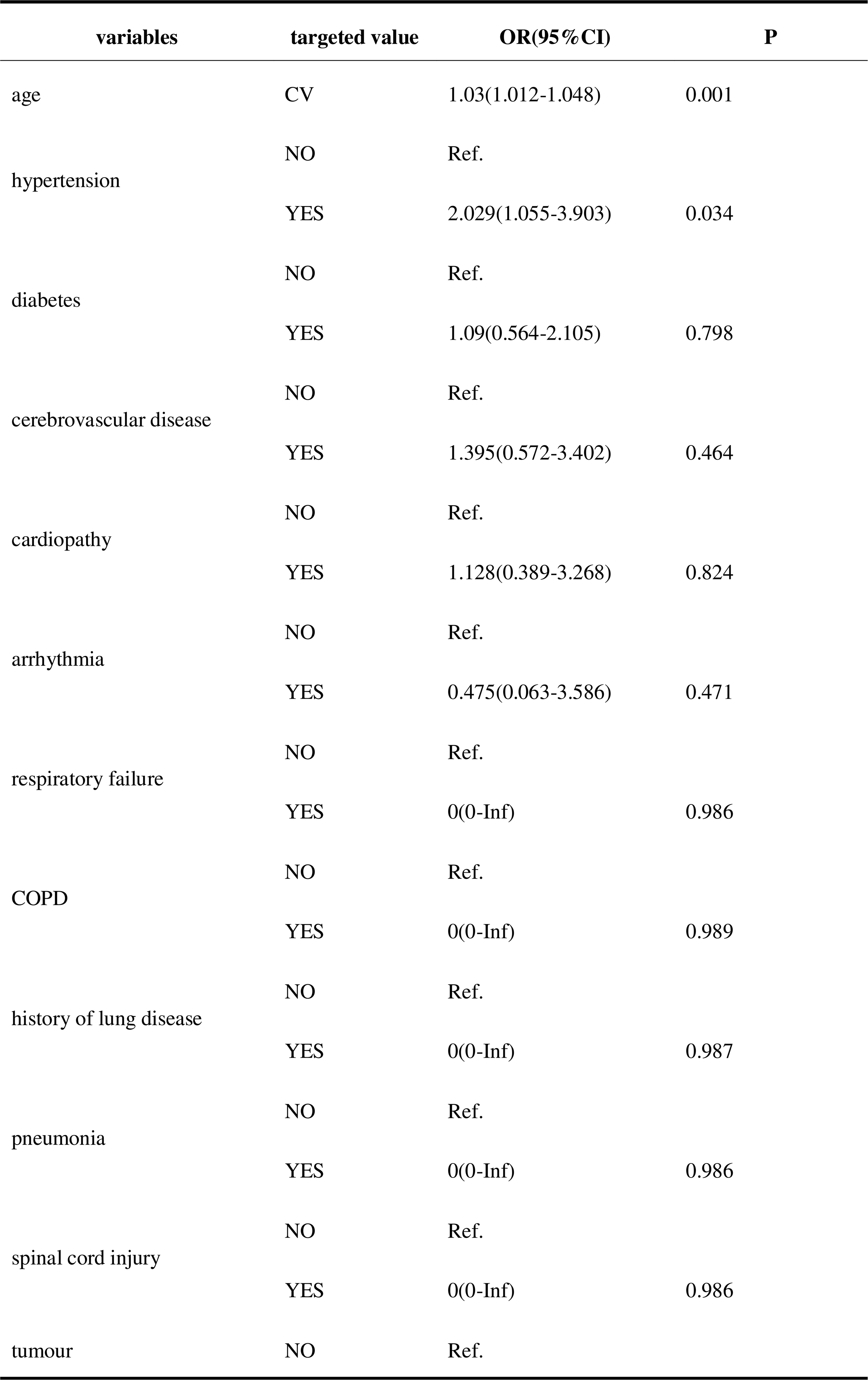
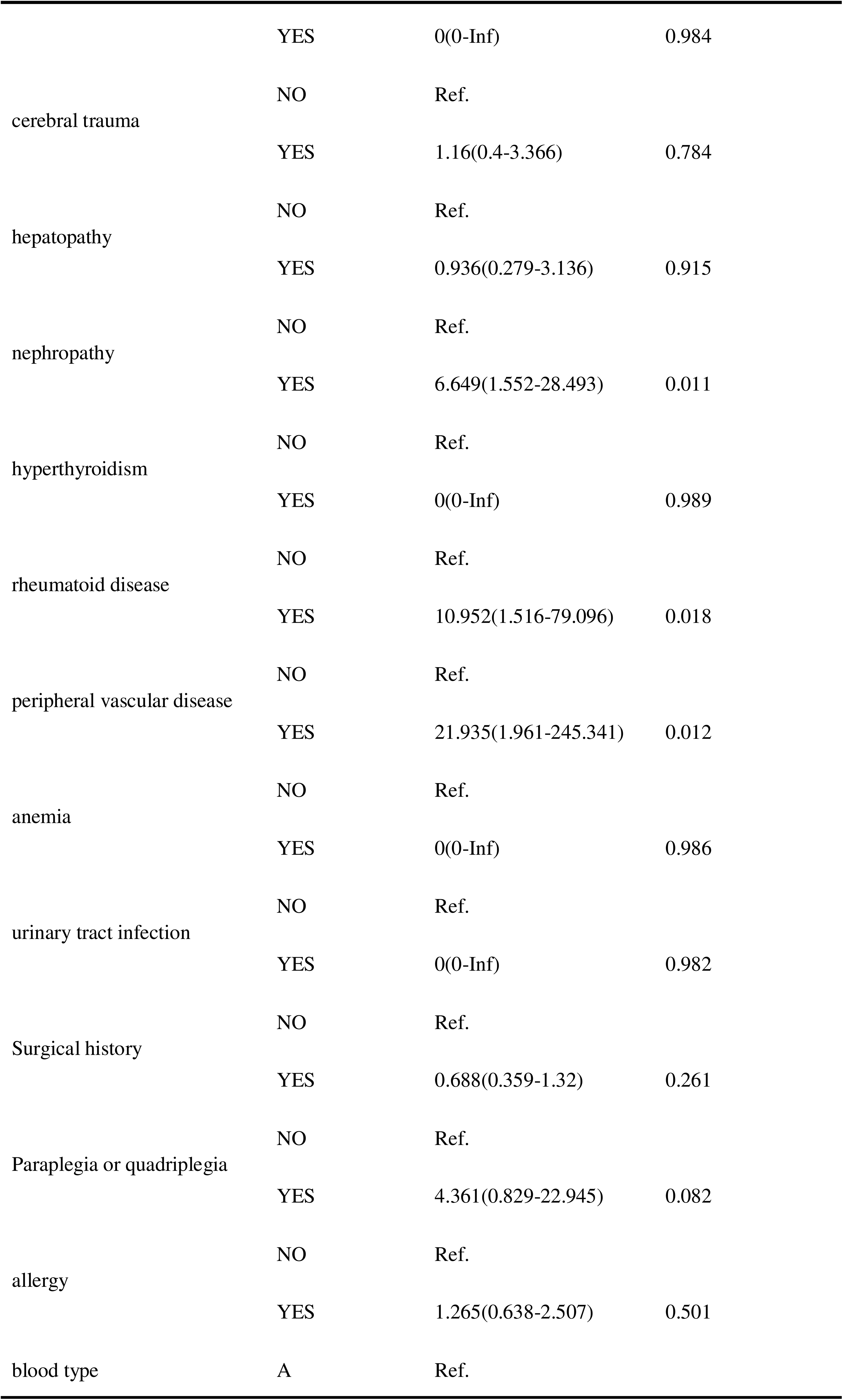
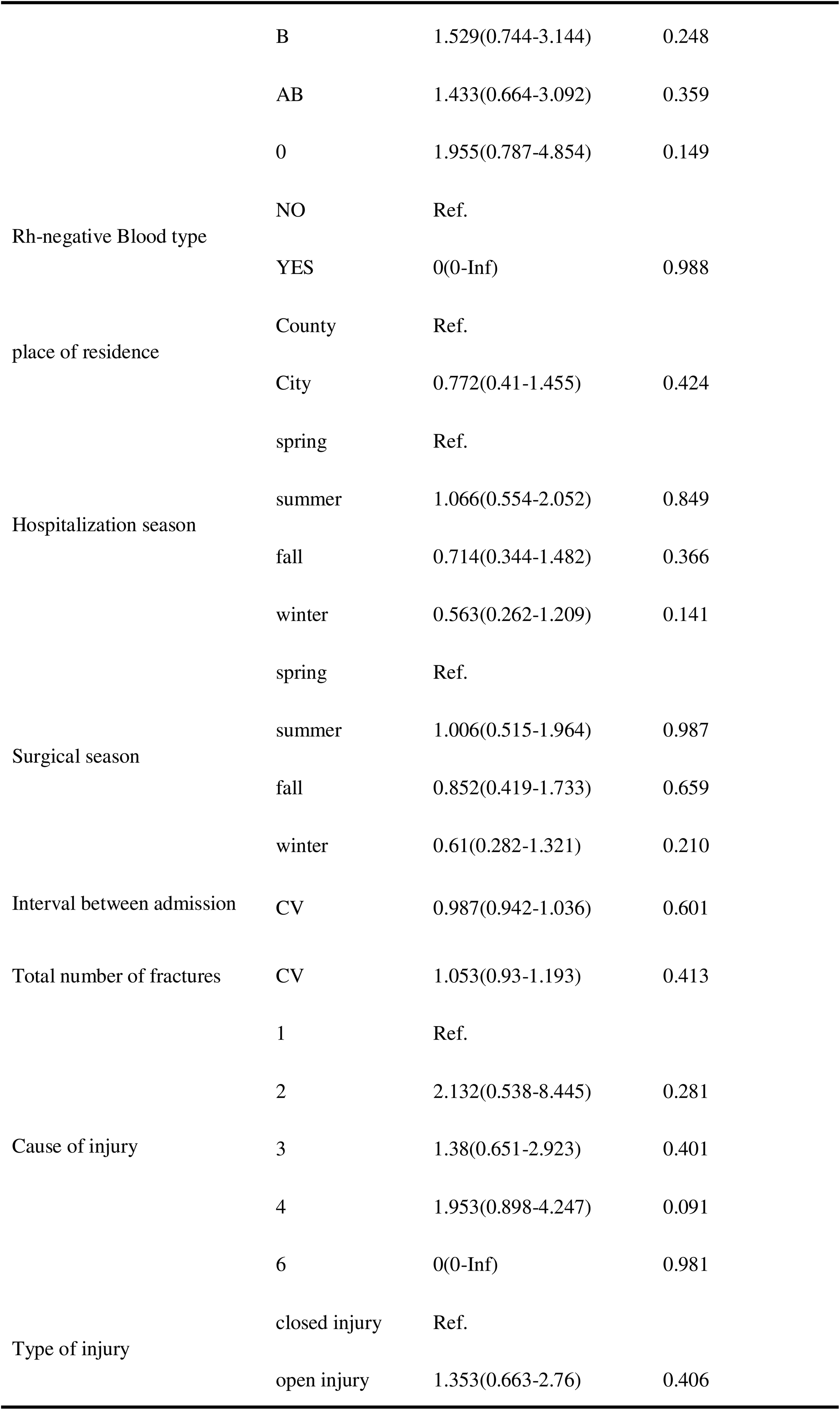
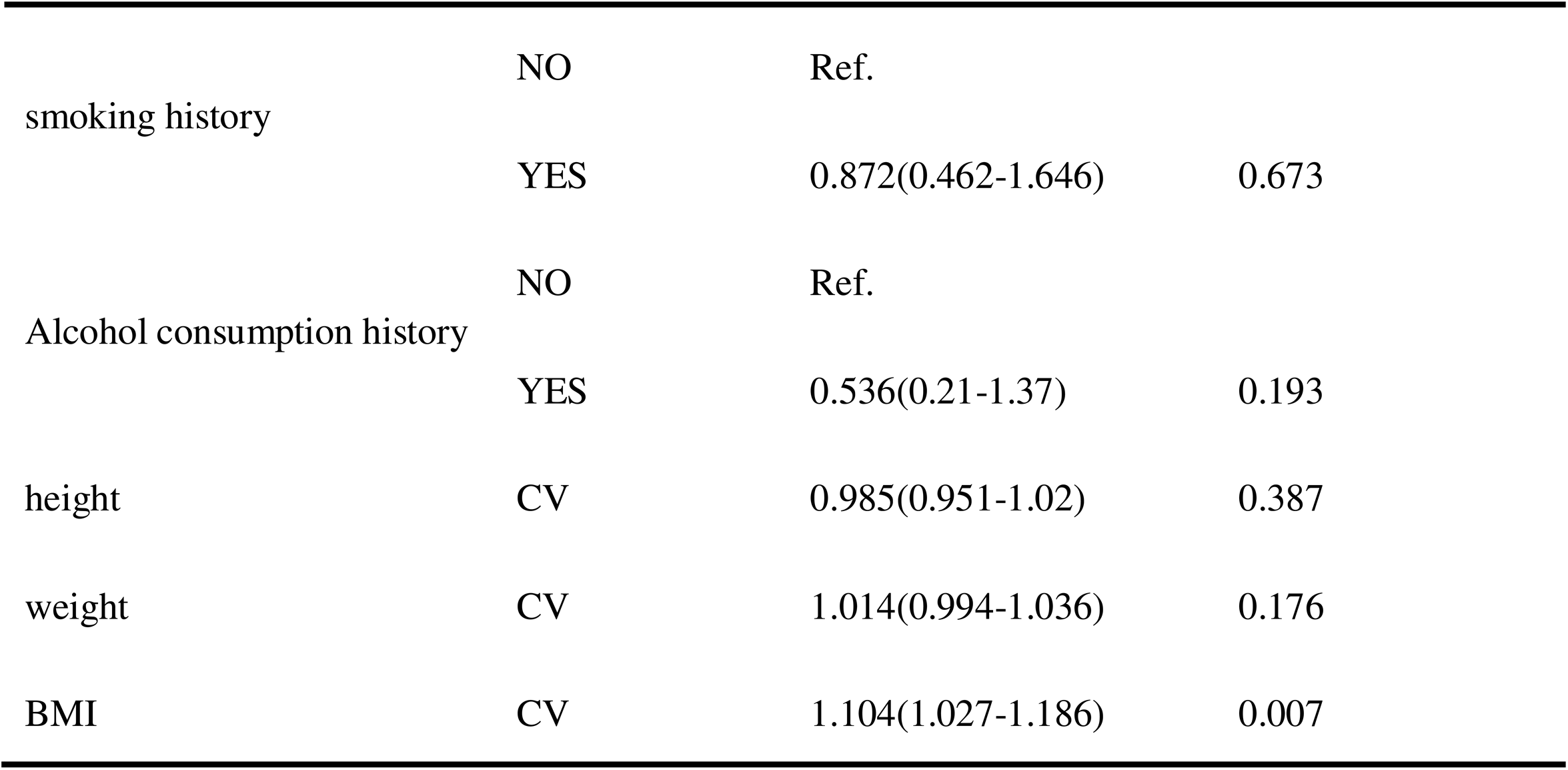
Univariate Analysis of General Information. Table 1: Univariate Analysis of General Information. Cause of injury (1 = Simple fall, 2 = Injury from electric bike or bicycle, 3 = Car accident injury, 4 = Injury from falling from a height, 5 = Injury from sharp objects, 6 = Other injuries)

The univariate logistic regression analysis of surgical and blood transfusion data (Table 2) identified several potential risk factors for postoperative thrombosis. These factors included surgical approach (fracture reduction), type of bone graft, duration of the operation, intraoperative blood loss, intraoperative blood transfusion, method of blood transfusion, and the administration of suspended red blood cells, plasma, and cryoprecipitate coagulation factors.

**Table 2:**
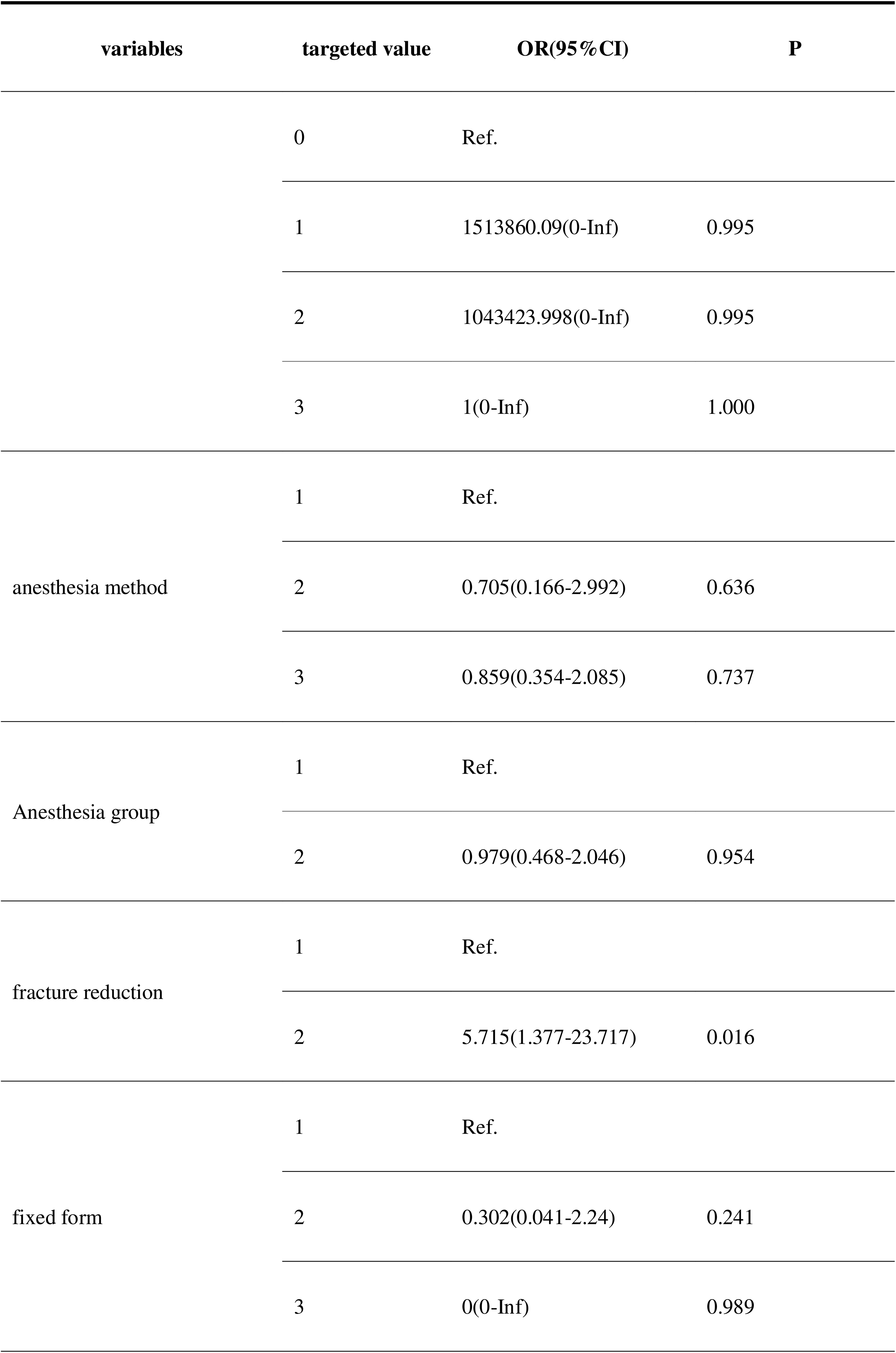
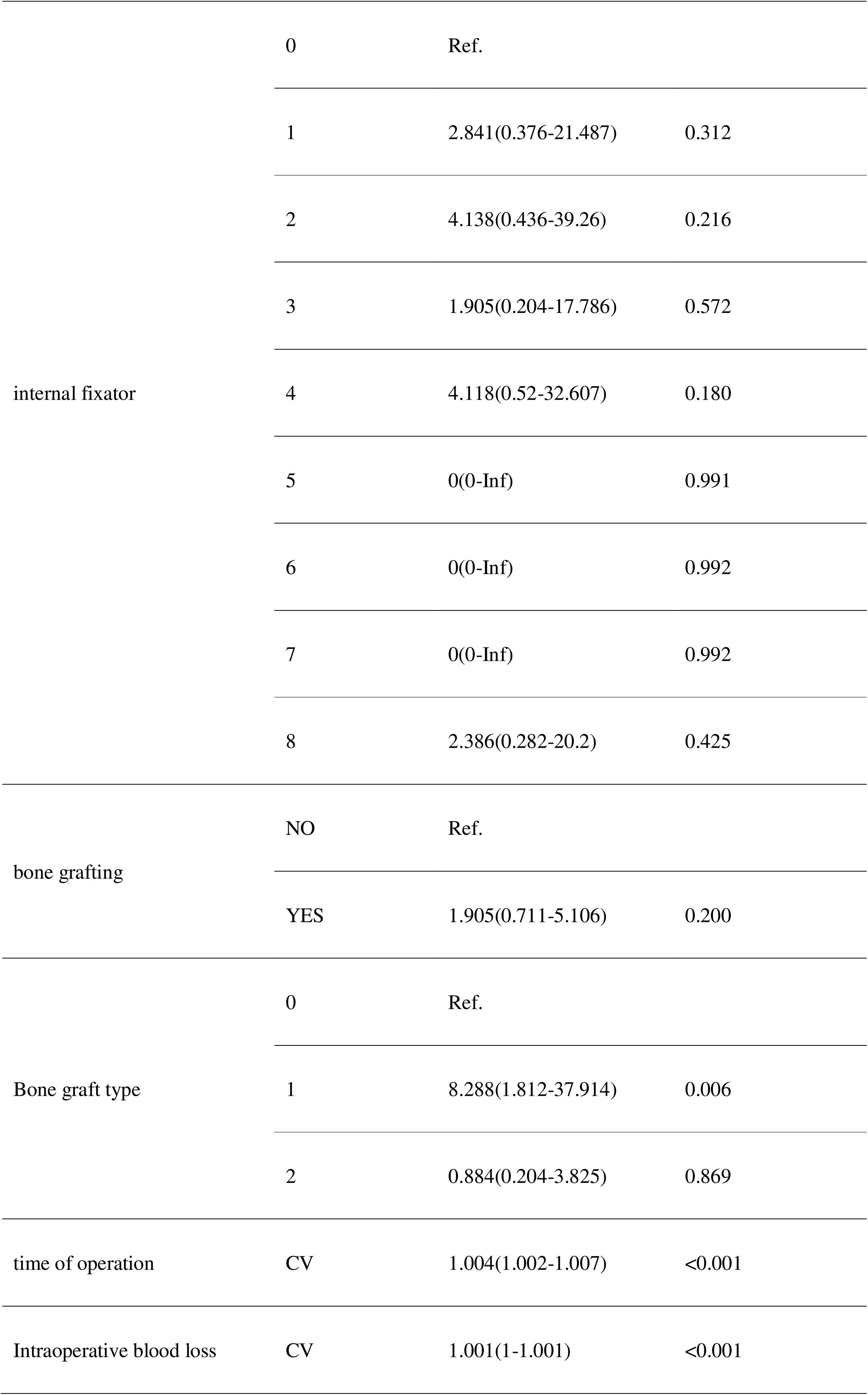
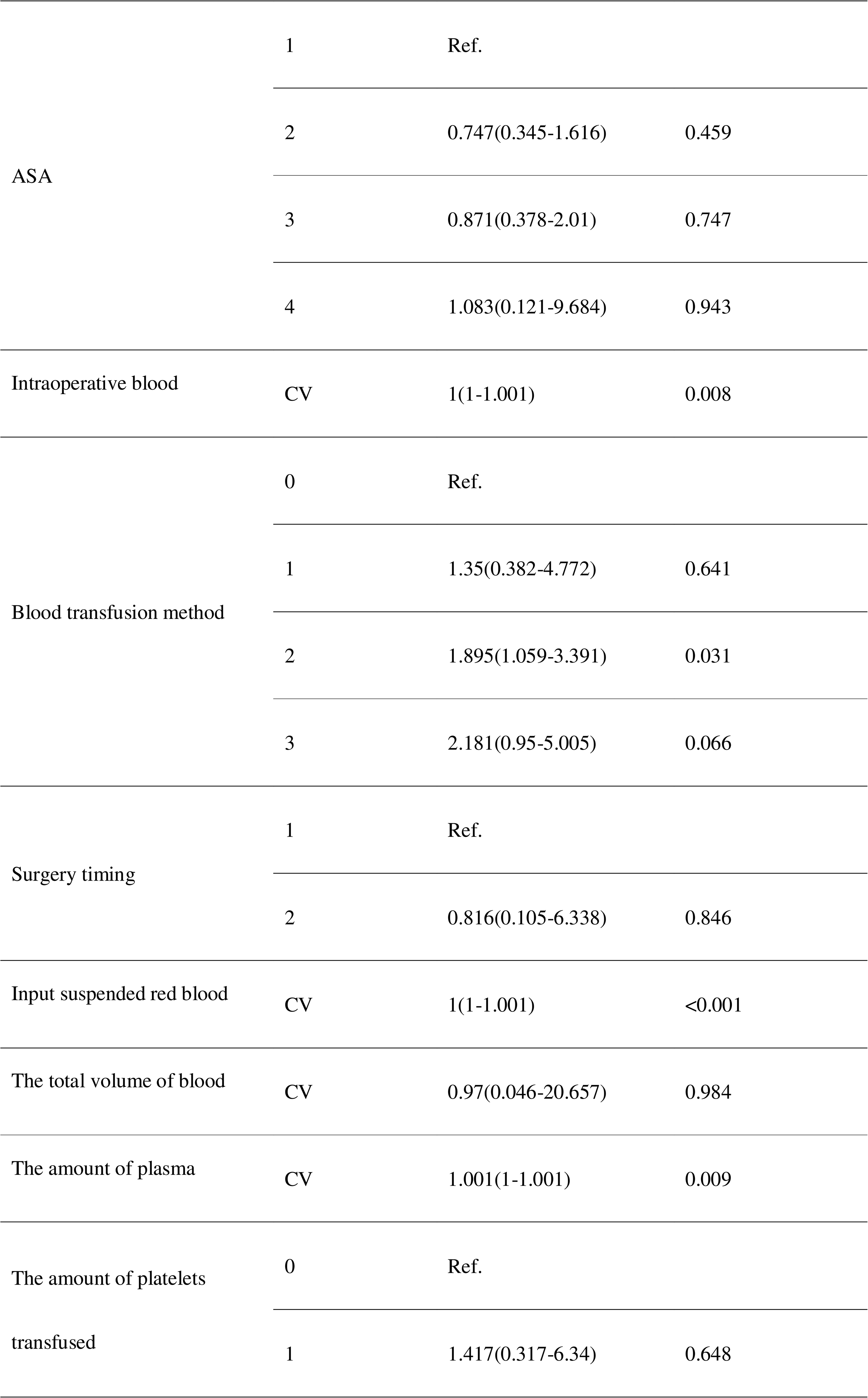
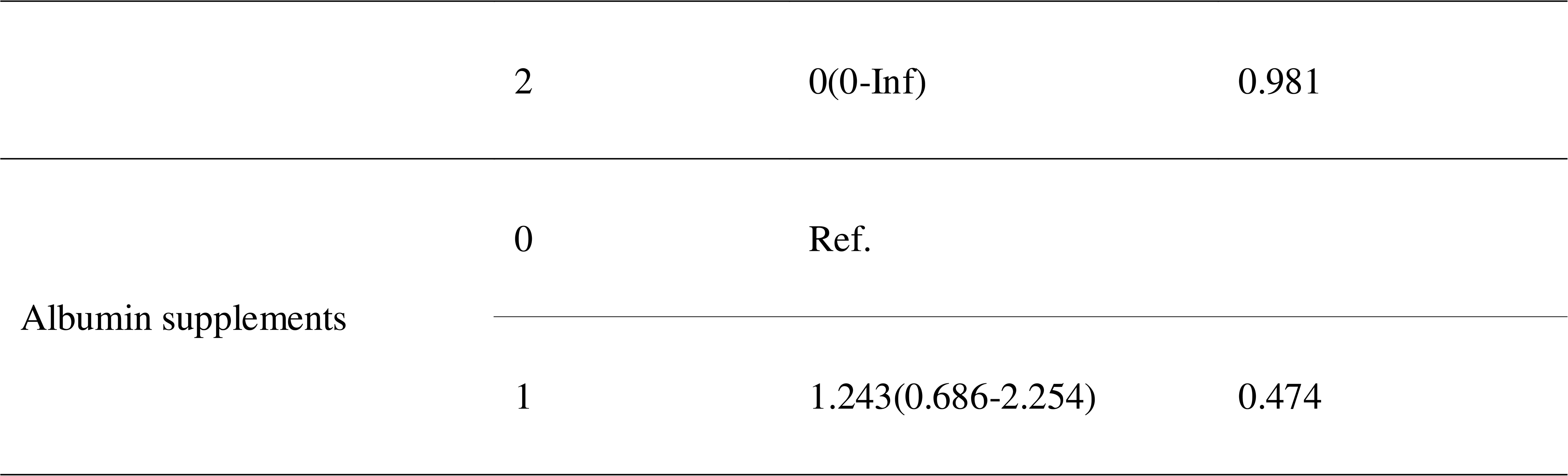
Univariate Analysis of Surgical and Blood Transfusion Information. Table 2: Univariate Analysis of Surgical and Blood Transfusion Information. Anesthesia method 1 = Local anesthesia (including brachial plexus, cervical plexus nerve block) 2 = Spinal anesthesia (including intradural, extradural anesthesia, and combined spinal-epidural anesthesia) 3 = General anesthesia; Anesthesia grouping: 1 local anesthesia; 2 general anesthesia; Reduction method 1 = Closed reduction 2 = Open reduction (or approach); Fixation method 1 = Internal fixation 2 = External fixation 3 = Conservative; Internal fixation device 0 = None 1 = Plate 2 = Intramedullary nail 3 = Kirschner wire 4 = Simple screw 5 = Total hip prosthesis 6 = Semi-hip prosthesis 7 = Rivet 8 = Others; Bone graft type 0 = No bone grafting 1 = Autologous bone 2 = Allogeneic bone; Blood transfusion method 0 = No blood transfusion 1 = Autologous blood transfusion 2 = Allogeneic blood transfusion 3 = Autologous + allogeneic blood transfusion; Surgery timing 1 = Daytime surgery 2 = Nighttime surgery

The results of the univariate logistic regression analysis, presented in Table 3 of the preoperative laboratory data, indicated that Gamma-Glutamyl Transferase (GGT), High-Density Lipoprotein Cholesterol (HDL-C), Apolipoprotein B (ApoB), Total Carbon Dioxide (TCO2), Glucose (GLU), Glasgow Admission Prognostic Score (GAP), Eosinophil Count (EOS), and Mean Platelet Volume (MPV) represented potential risk factors for postoperative thrombosis.

**Table 3:**
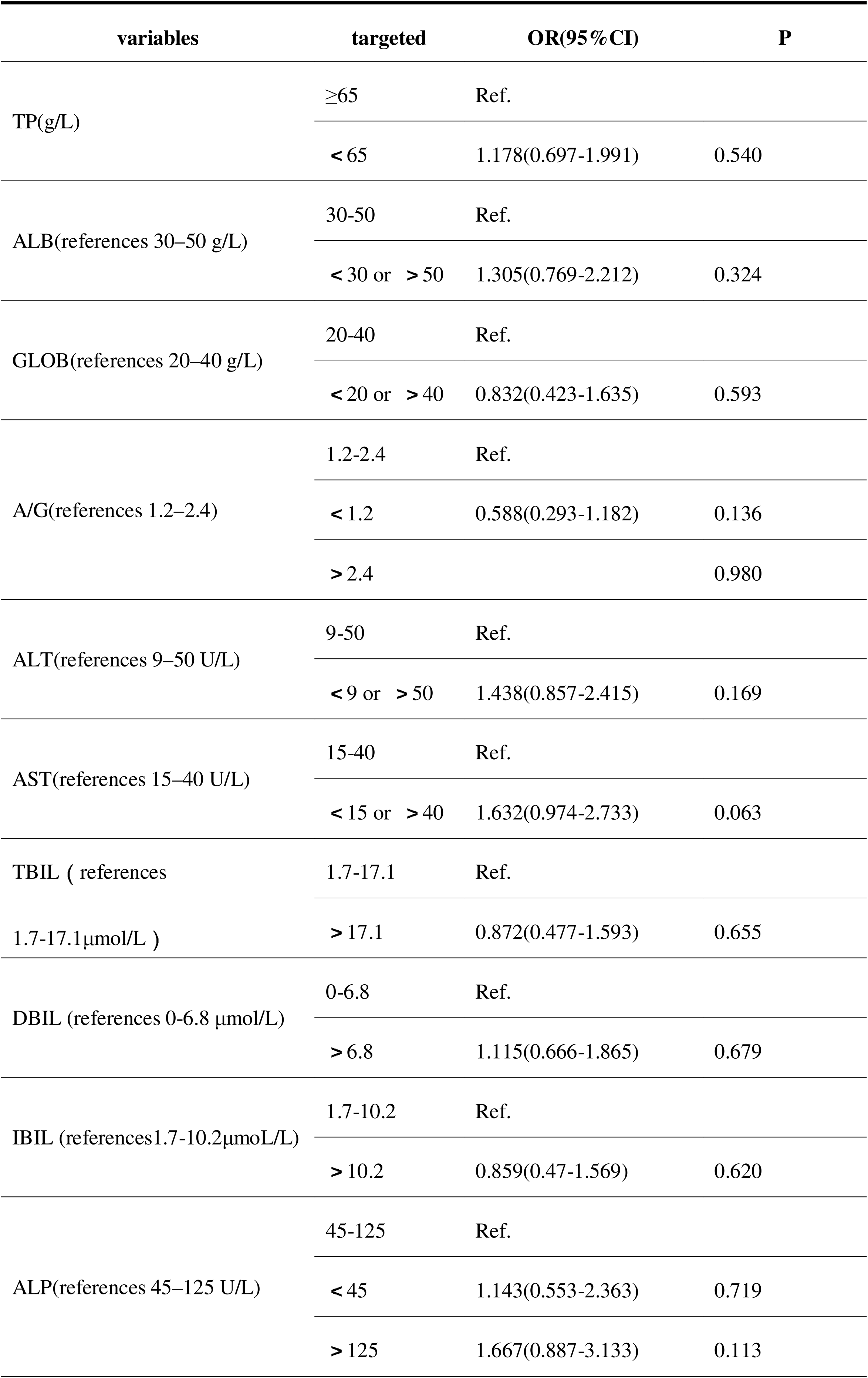
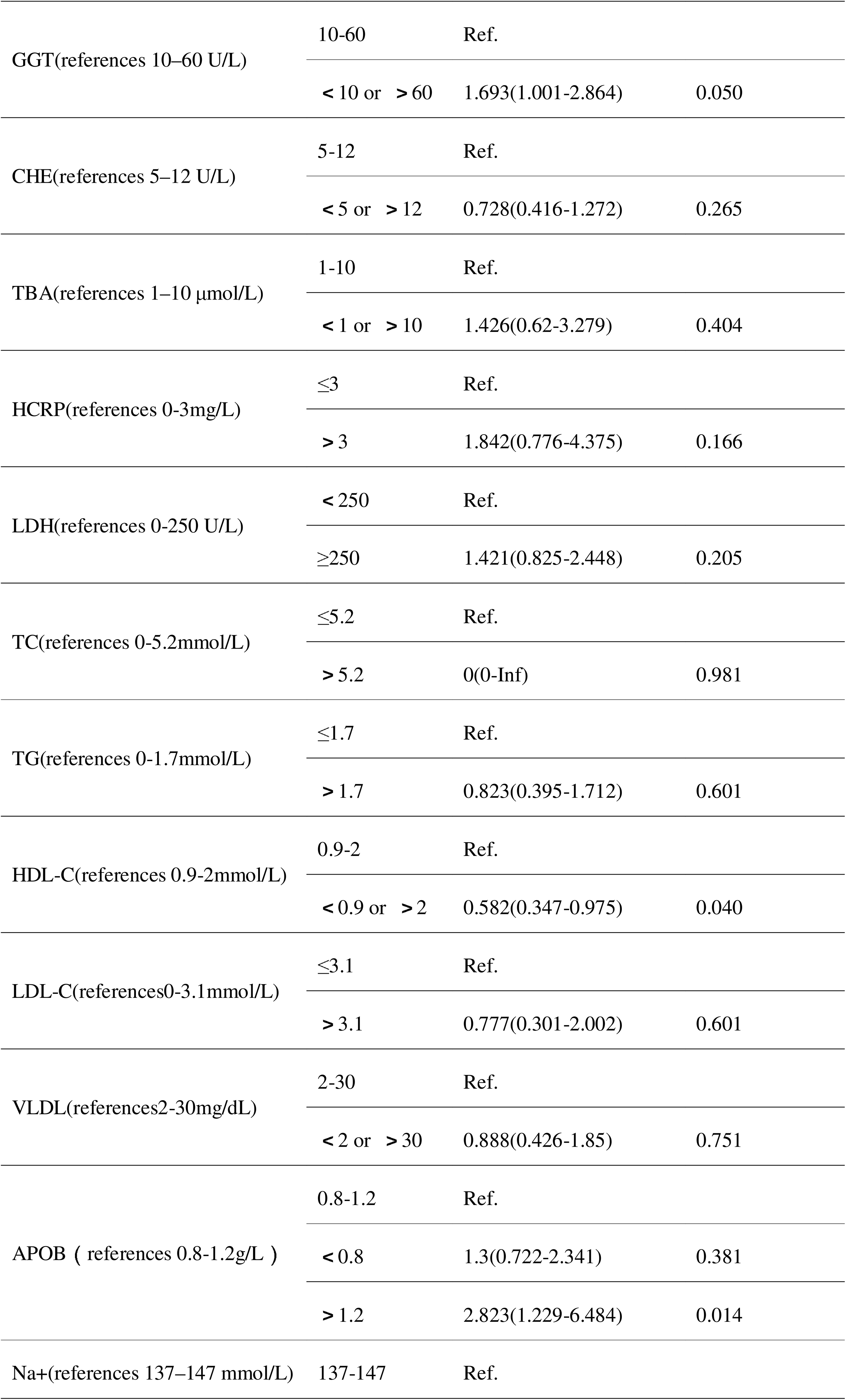
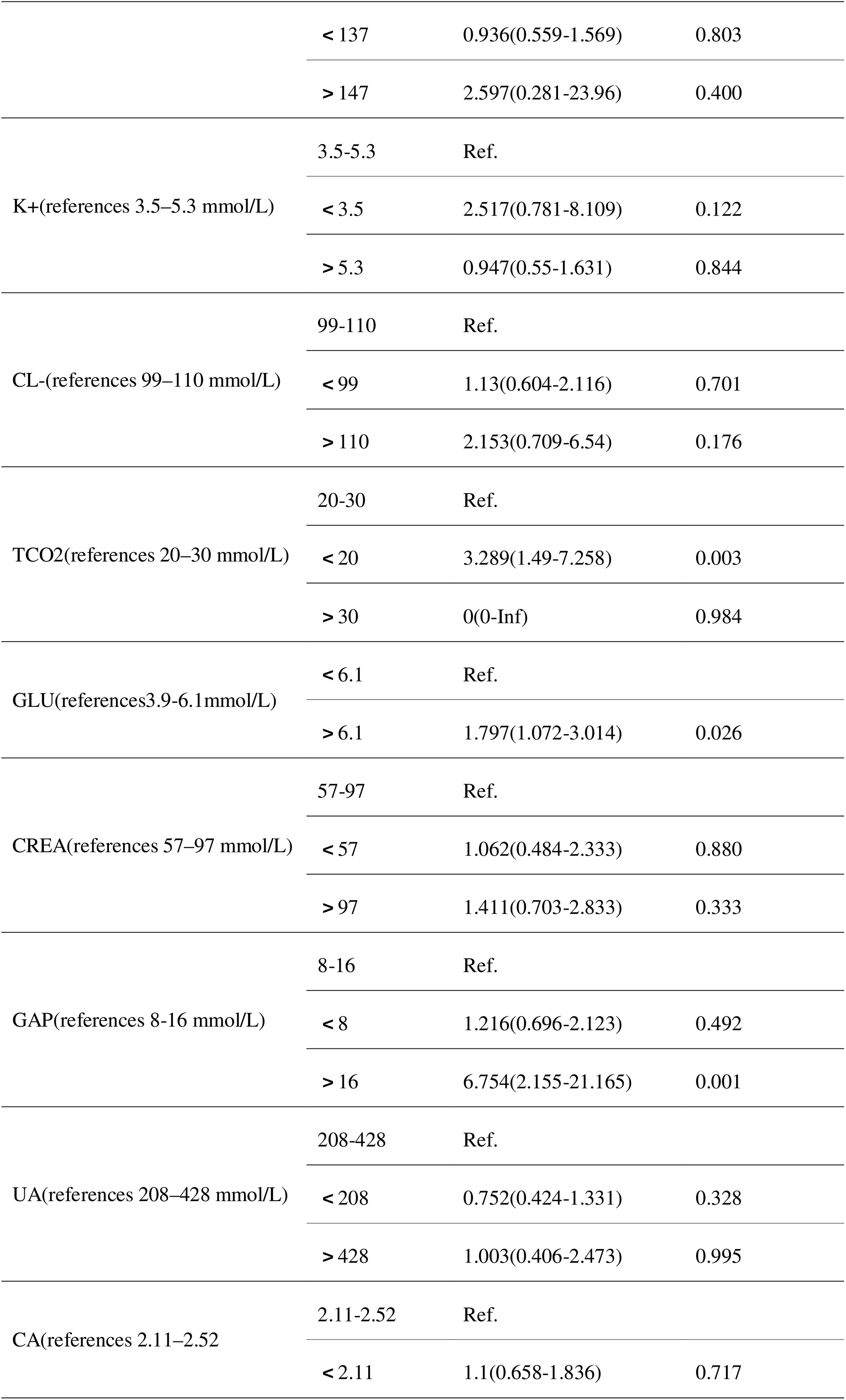
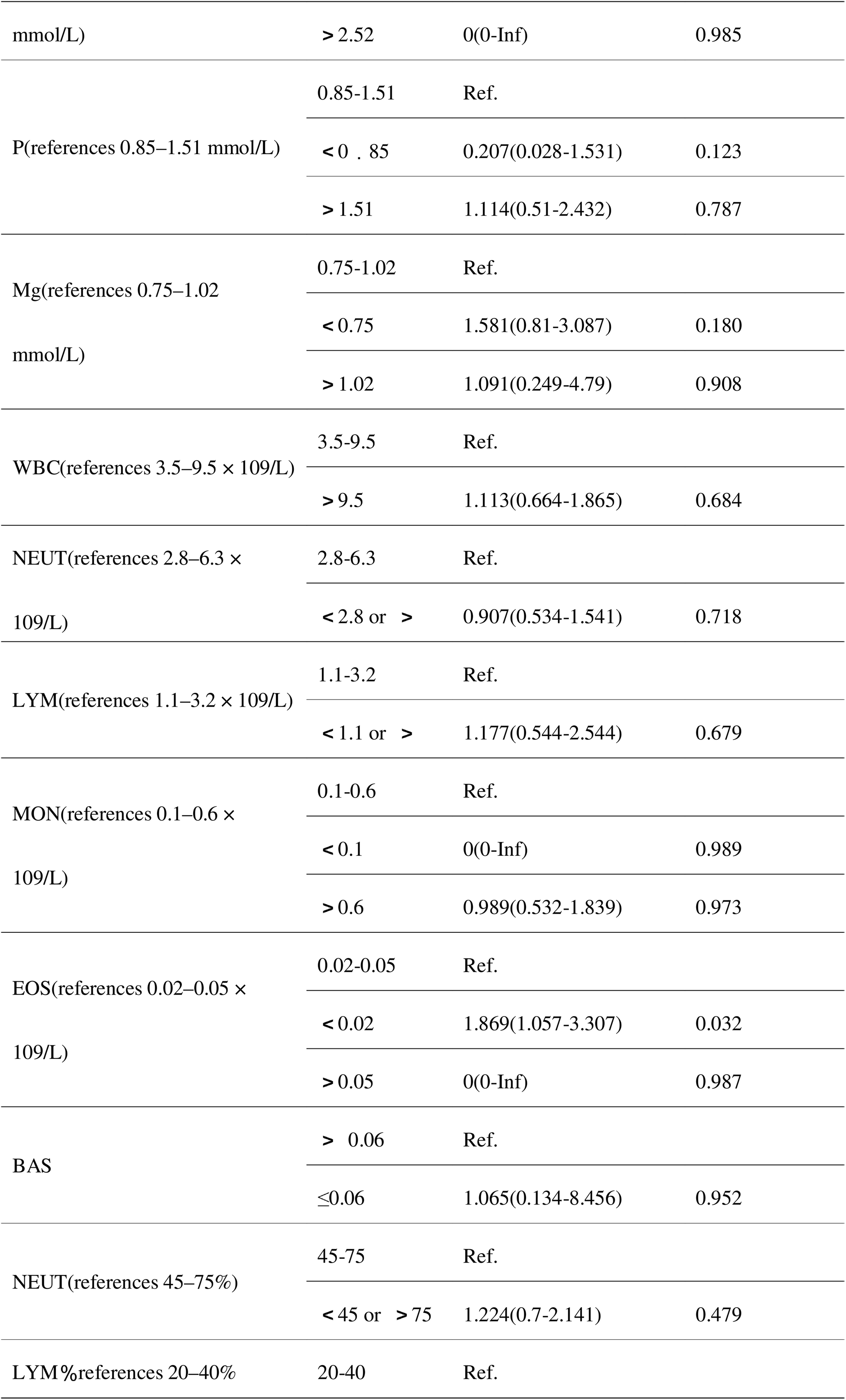
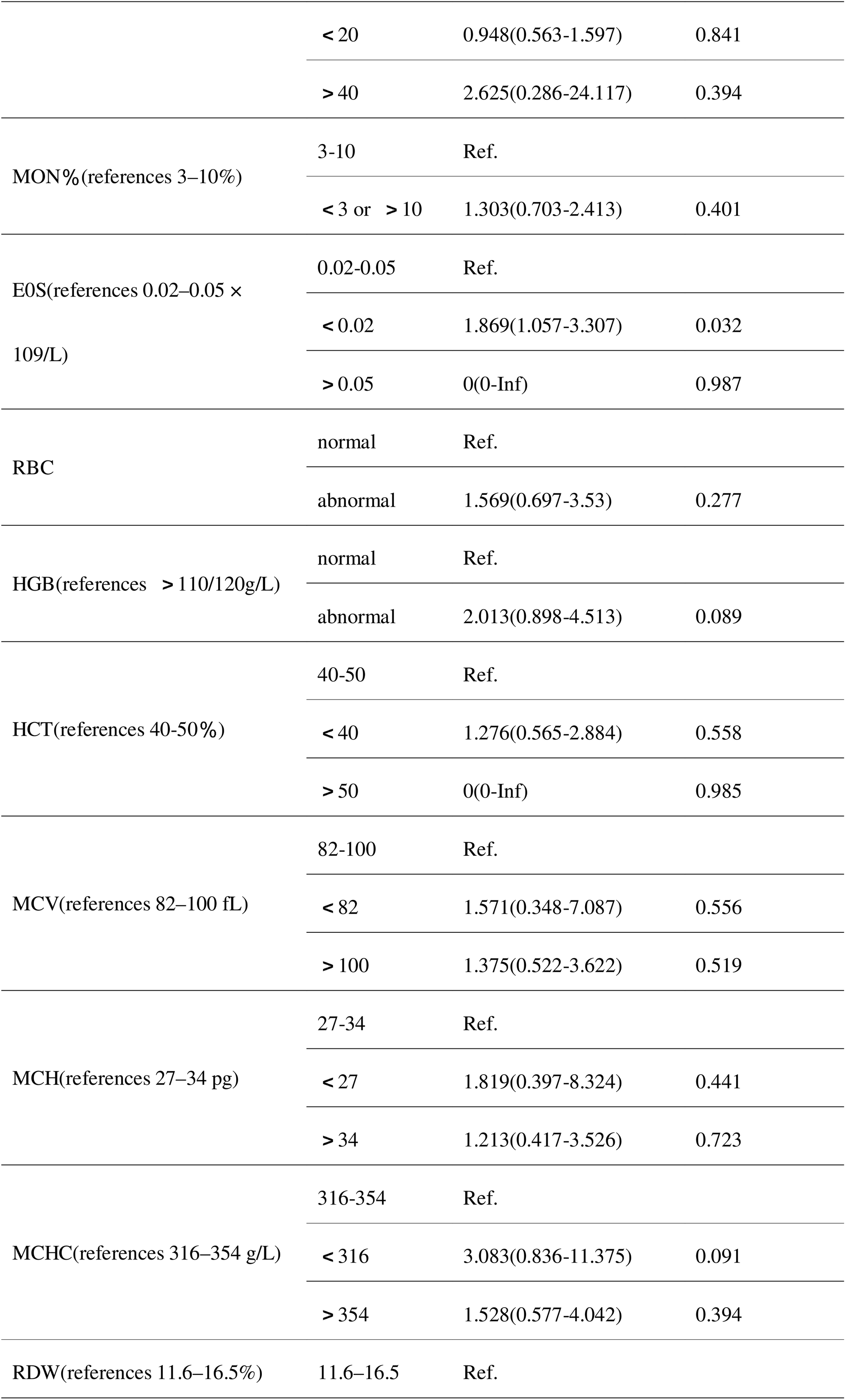
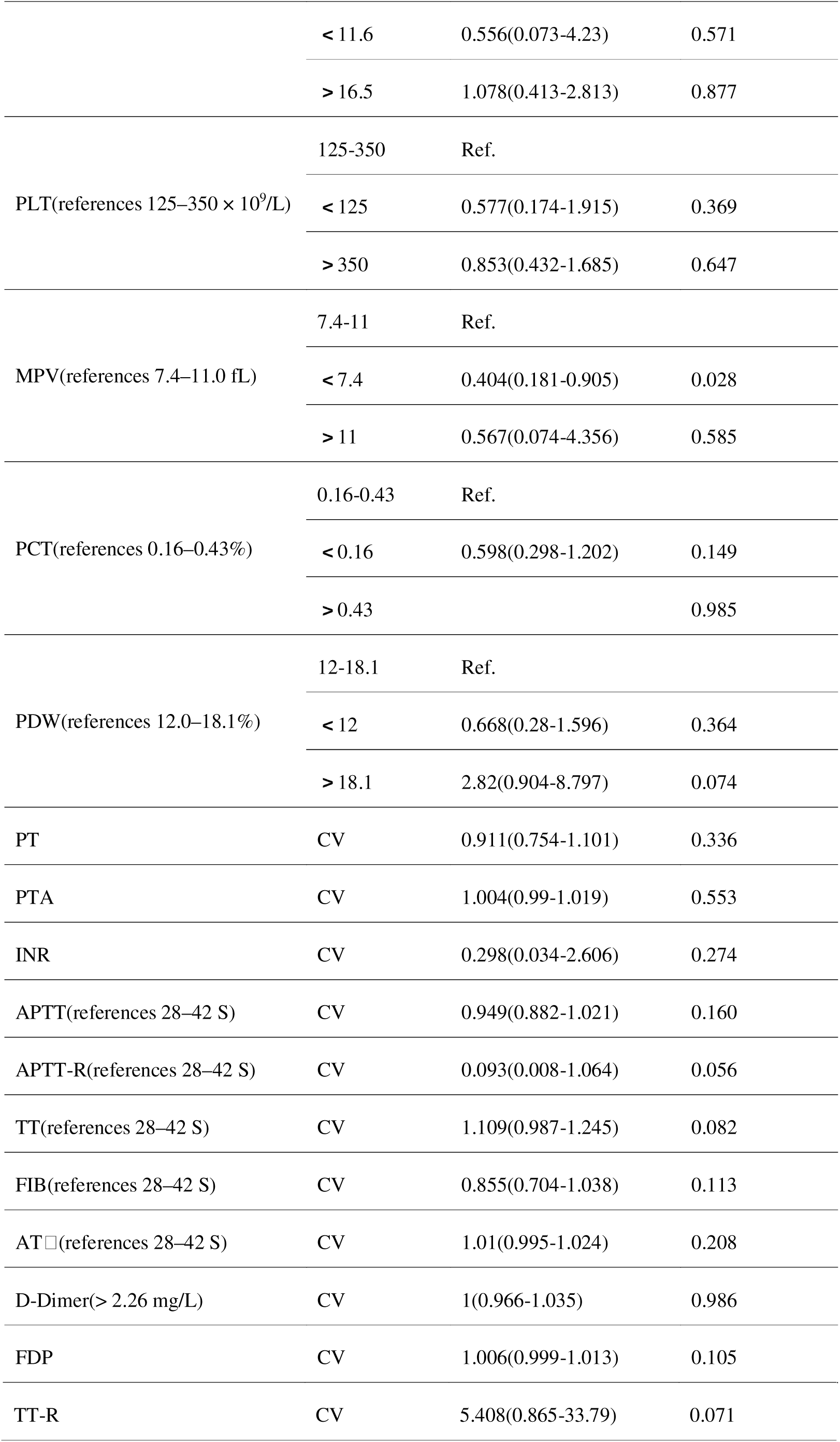
Univariate Analysis of Laboratory Information. Table 3: Univariate Analysis of Laboratory Information. Comparison of preoperative laboratory indicators between the two groups of patients. TP - total protein, ALB - albumin, GLOB - globulin, A/G value (albumin/globulin), ALT - alanine aminotransferase, AST - aspartate aminotransferase, TBIL - total bilirubin, DBIL - direct bilirubin, IBIL - indirect bilirubin, ALP - alkaline phosphatase, GGT - γ-glutamyl transpeptidase, CHE - phenol red enzyme, TBA - total bile acid, HCRP - high-sensitivity C-reactive protein, LDH - lactate dehydrogenase, CREA - creatinine, UA - uric acid, CA - calcium, P - phosphorus, Mg - magnesium, BNP - brain natriuretic peptide, WBC - white blood cells, NEUT - neutrophils, LYM - lymphocytes, MON - monocytes, EOS - eosinophils, BAS - basophils, RBC - red blood cells. Reference range: for females, 3.5–5.0 × 10^12/L; for males, 4.0–5.5 × 10^12/L. HGB - hemoglobin, reference range: for females, 110–150 g/L; for males, 120–160 g/L. HCT - hematocrit, 40–50%, MCV - mean cell volume, MCH - mean corpuscular hemoglobin content, MCHC - mean corpuscular hemoglobin concentration, RDW - red cell distribution width, PLT - platelets, 100–300 × 10^9/L, MPV - mean platelet volume. Platelet volume, PCT (procalcitonin), PDW (platelet distribution width), PT (prothrombin time), PTA (prothrombin activity), INR (international normalized ratio), APTT (activated partial thromboplastin time), APTT-R (ratio of activated partial thromboplastin time), TT (thrombin time), TT-R (thrombin ratio), FIB (fibrinogen), ATIII (antithrombin 3).

Further analysis evaluated systemic inflammatory markers, including the Systemic Immune-Inflammation Index (SII), Neutrophil-to-Lymphocyte Ratio (NLR), and Platelet-to-Lymphocyte Ratio (PLR). However, none of these indicators demonstrated significant differences, as presented in Table 4.

**Table 4:**
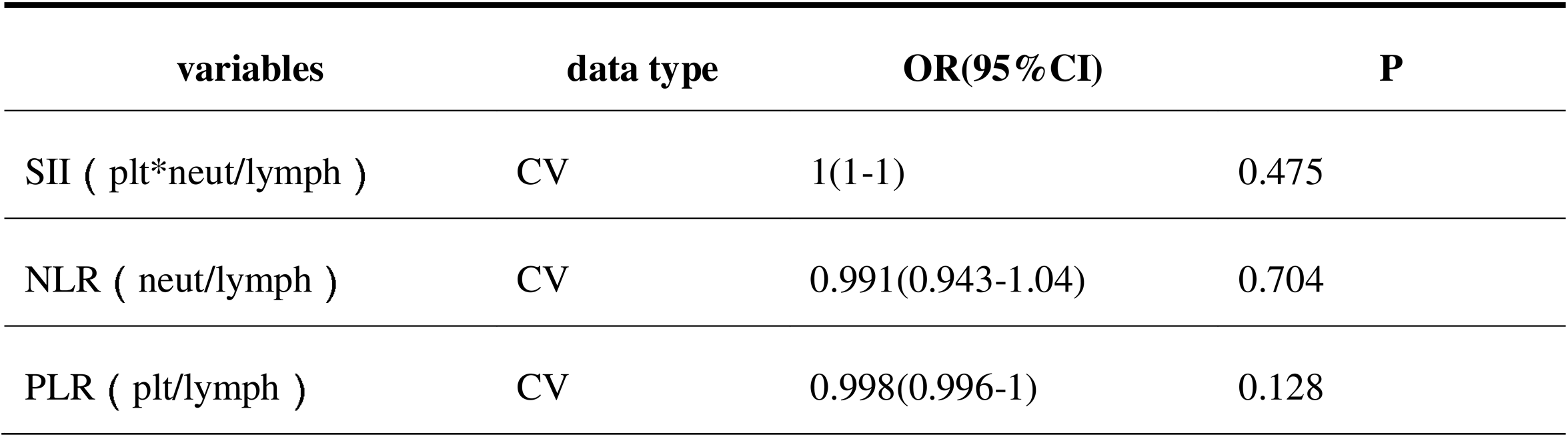
Univariate Analysis of Validation Index Scores.

### 2. LASSO regression for feature selection

LASSO regression was employed to further identify the factors influencing postoperative thrombosis. To mitigate collinearity in the data analysis, LASSO regression was used to select variables with *P*-values < 0.05 from the univariate analysis. As λ increased, the penalty also increased, leading to the gradual shrinkage of the regression coefficients of the independent variables towards zero. This regularization term effectively reduced model overfitting. Figures 1 and 2 illustrate the variable selection process using LASSO regression and the impact of cross-validation on overfitting. In Figure 1, each curve represents the trajectory of a variable’s coefficient, with the ordinate indicating the coefficient value and the abscissa showing the parameter λ, which controls the regularization intensity. As λ varied, the coefficients of the variables progressively approached zero, ultimately retaining only those variables with non-zero coefficients. Figure 2 emonstrates the determination of the optimal λ value via 10-fold cross-validation, indicated by a vertical dotted line. The left dotted line illustrated that when Lambda was set to Lambda.min, 18 variables were selected, whereas the right dotted line demonstrated that when Lambda was set to Lambda.1se, 13 variables were selected. In this study, Lambda.1se (i.e., Lambda = 0.01641399) was selected as the optimal penalty coefficient for the model. Following the LASSO regression analysis, variables with non-zero regression coefficients were identified, namely age, nephropathy, peripheral vascular disease, BMI, fracture reduction, operation time, intraoperative blood loss, deerythrocytized RBC, HDL-C group, ApoB group, GLU group, GAP group, and MPV group, as presented in Table 5.

**Figure 1:**
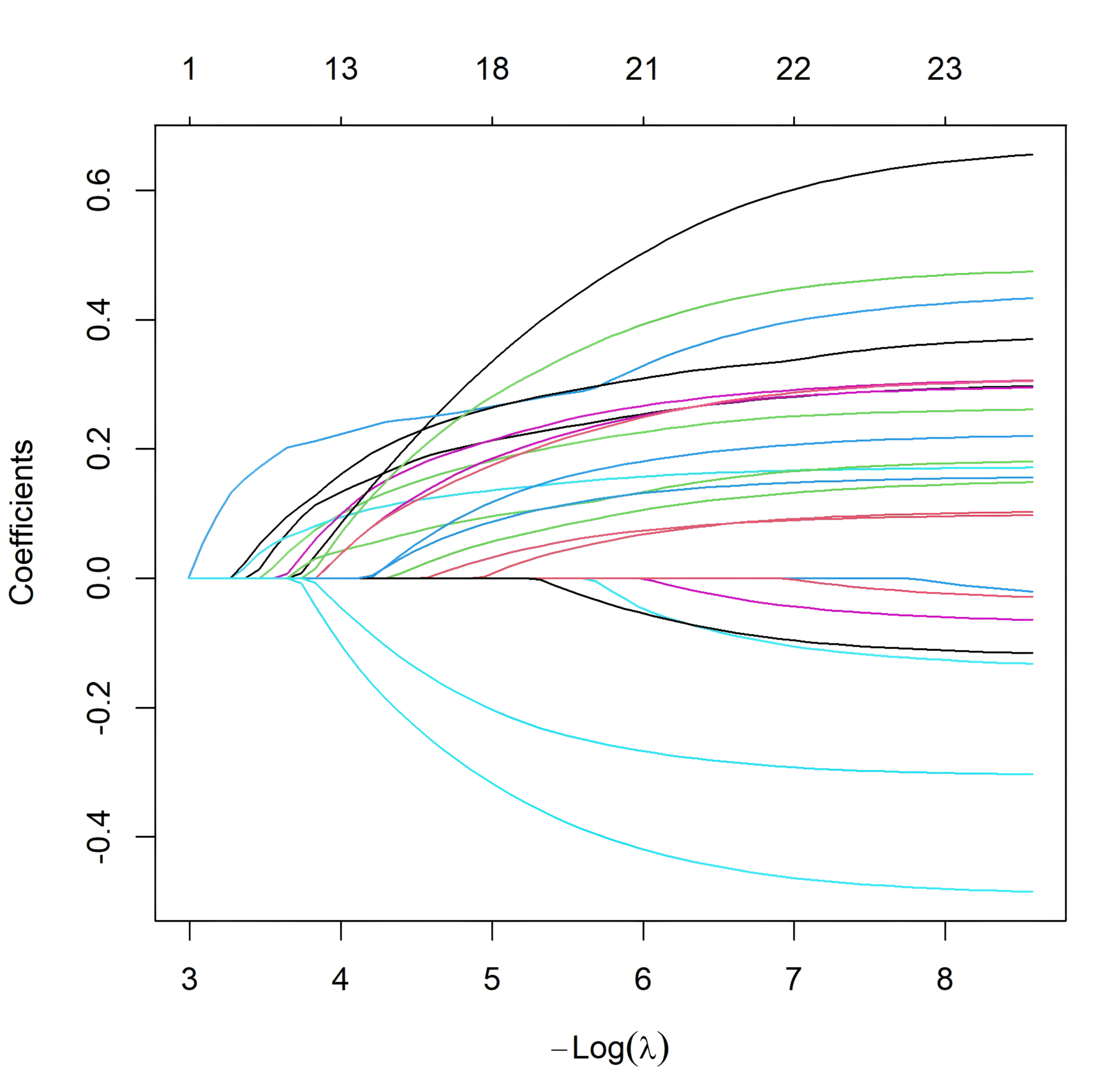
Feature selection process using Least Absolute Shrinkage and Selection Operator (LASSO) regression.

**Figure 2:**
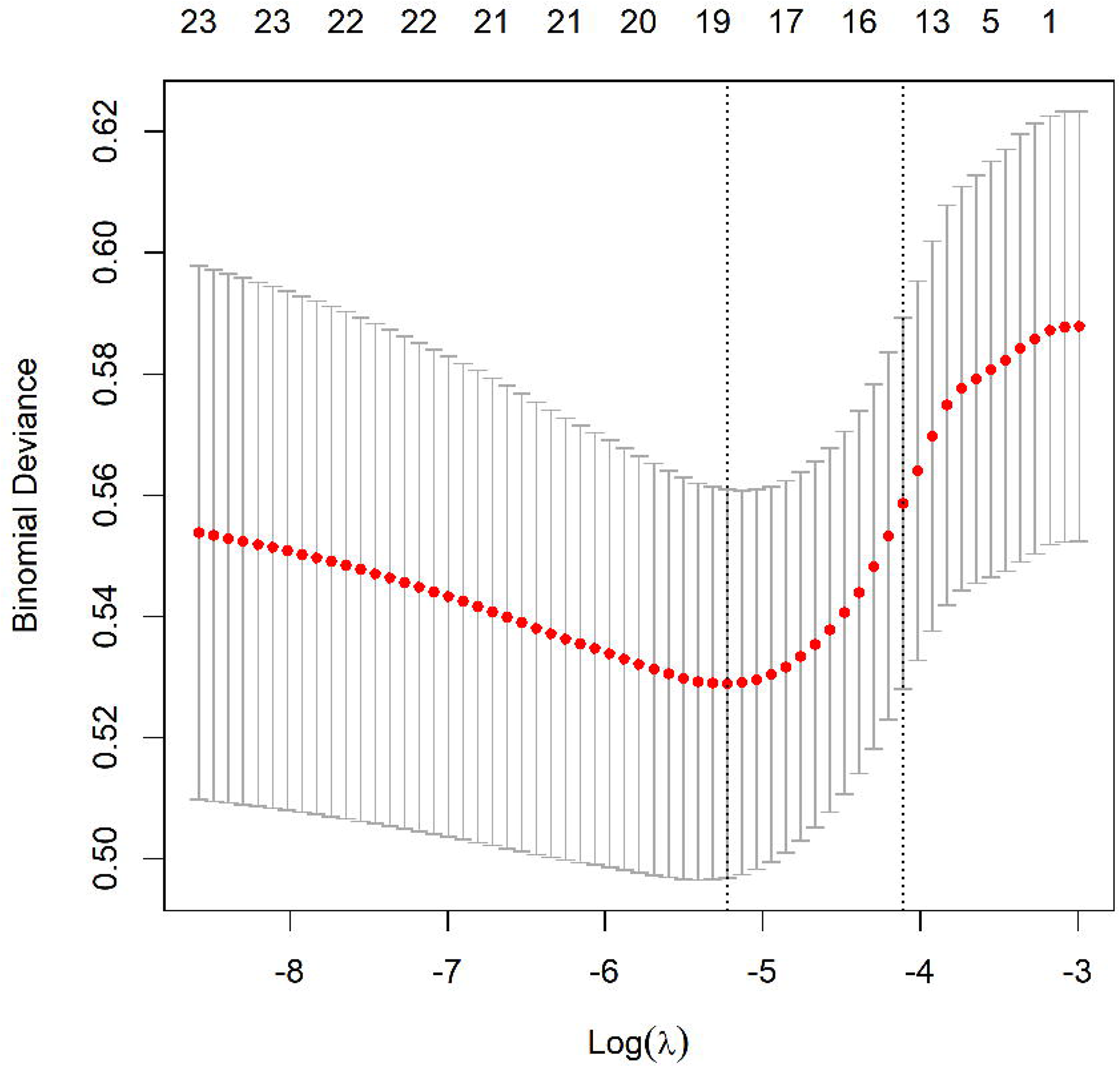
Graph of the Optimal *λ* Selection for Cross-Validation

**Table 5:**
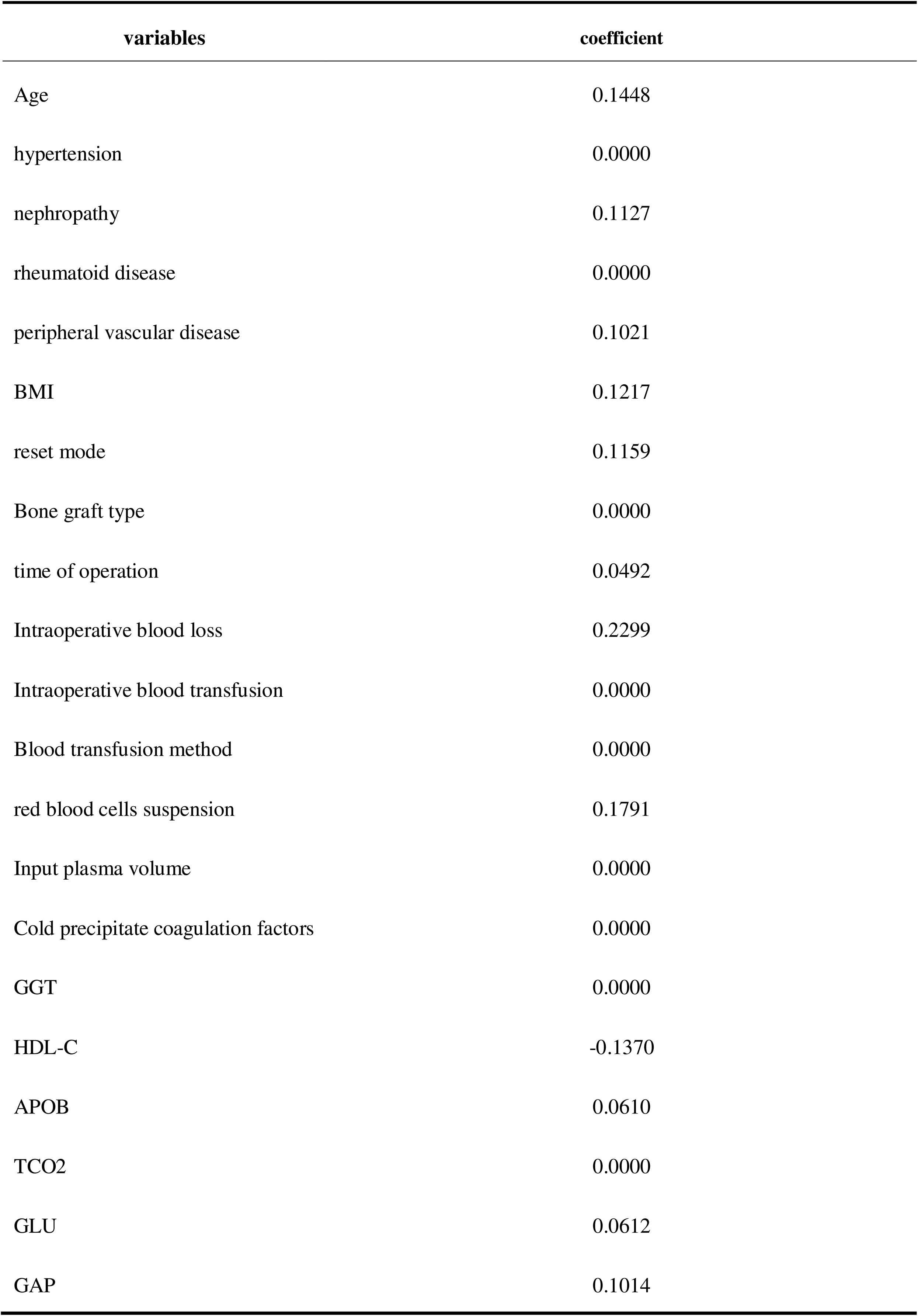
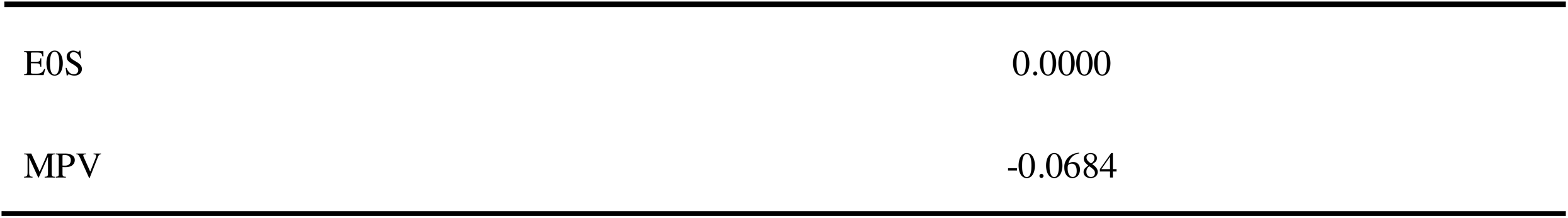
Lasso Result Coefficient Table.

### 3. Multivariate Logistic Regression Analysis

Building on the nine variables identified through univariate analysis and LASSO regression, a multivariate logistic regression analysis was conducted to identify independent predictors of postoperative thrombosis. The assignment outcomes for the independent variables are presented in the table below. Our analysis identified age, nephropathy, BMI, fracture reduction,intraoperative blood loss, administration of red blood cell suspension, HDL-C, ApoB, GAP, and MPV as independent risk factors for blood transfusion. The detailed results of the regression analysis are presented in Table 6.

**Table 6:**
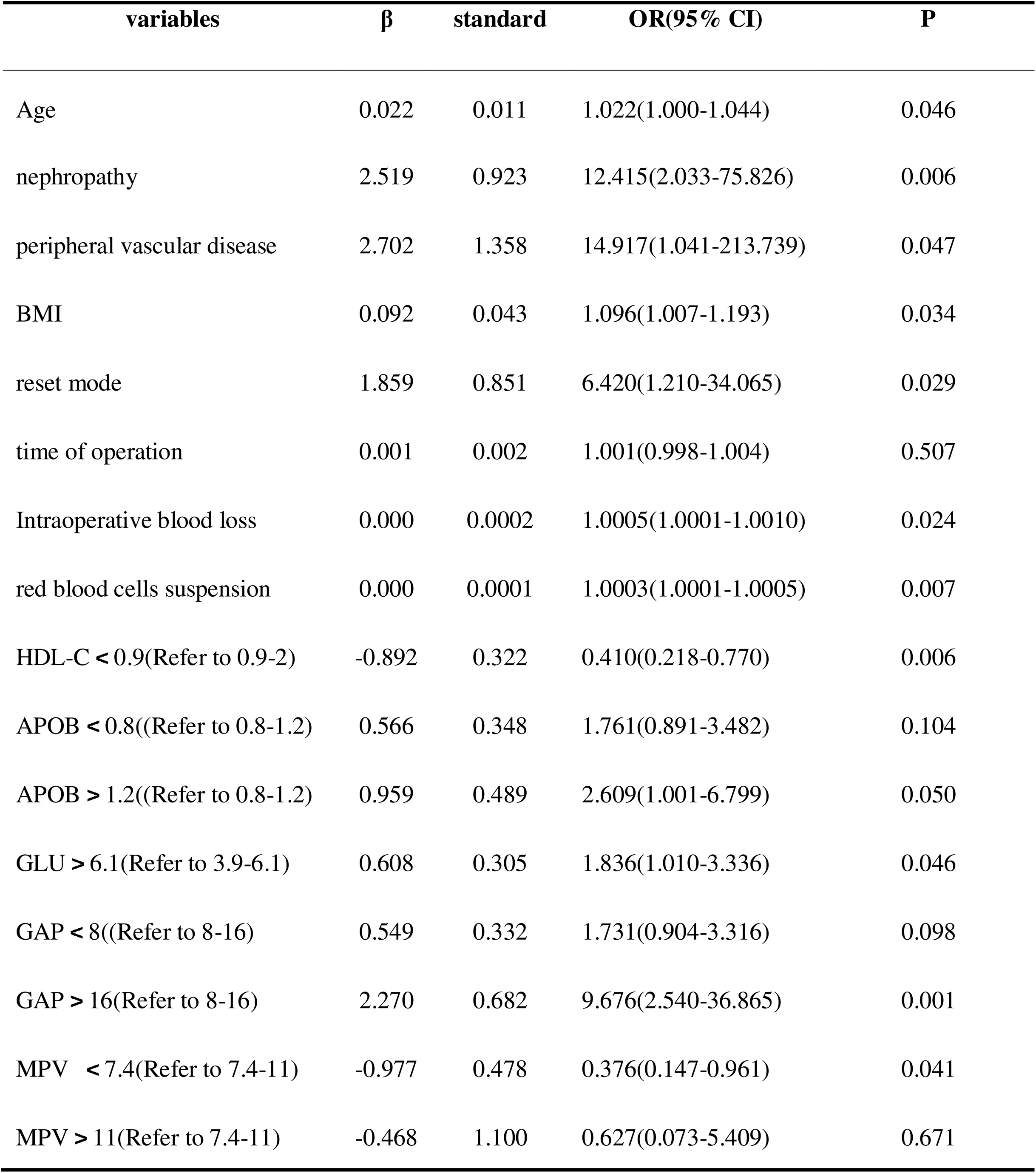
Independent Predictors of Postoperative Thrombosis in Multivariate Logistic Regression Analysis.

#### Establishment and Analysis of Postoperative PNO-DVT Prediction Model Based on Machine Learning Algorithms

##### 1. Establishment of the prediction model and performance evaluation

The twelve significant features identified from the preceding univariate analysis were selected as the dataset for the final model construction. The dataset was randomly split into a training set (70%) and a test set (30%). Given that the ratio of positive to negative samples was approximately 1:10, there was a notable imbalance. To mitigate the risk of the model being biased towards the negative samples during prediction, which could result in an unacceptably low recall rate, sampling was conducted on the training set; however, no processing was applied to the test set. This approach ensured that the model evaluation more accurately reflected the model’s predictive capabilities. Six machine learning algorithms, including logistic regression, SVM, random forest, XGBoost, LightGBM, and AdaBoost, were employed to construct six distinct models. Among these, the XGBoost algorithm achieved the highest AUC value of 0.8633, indicating superior predictive performance for postoperative thrombosis within this dataset (Table 7). Consequently, XGBoost was determined to be the optimal model( Figure 3). Supplementary Figure 1 illustrates the feature importance rankings for the random forest, XGBoost, LightGBM, and AdaBoost models. To ensure the robustness and generalizability of model performance, we applied fiveLJfold crossLJvalidation within the training set, whereby the data were randomly partitioned into five equal subsets; in each iteration, four subsets were used for model training and the remaining subset for validation, and the final performance was obtained by averaging the results across all five folds (Supplementary File S1).

**Figure 3:**
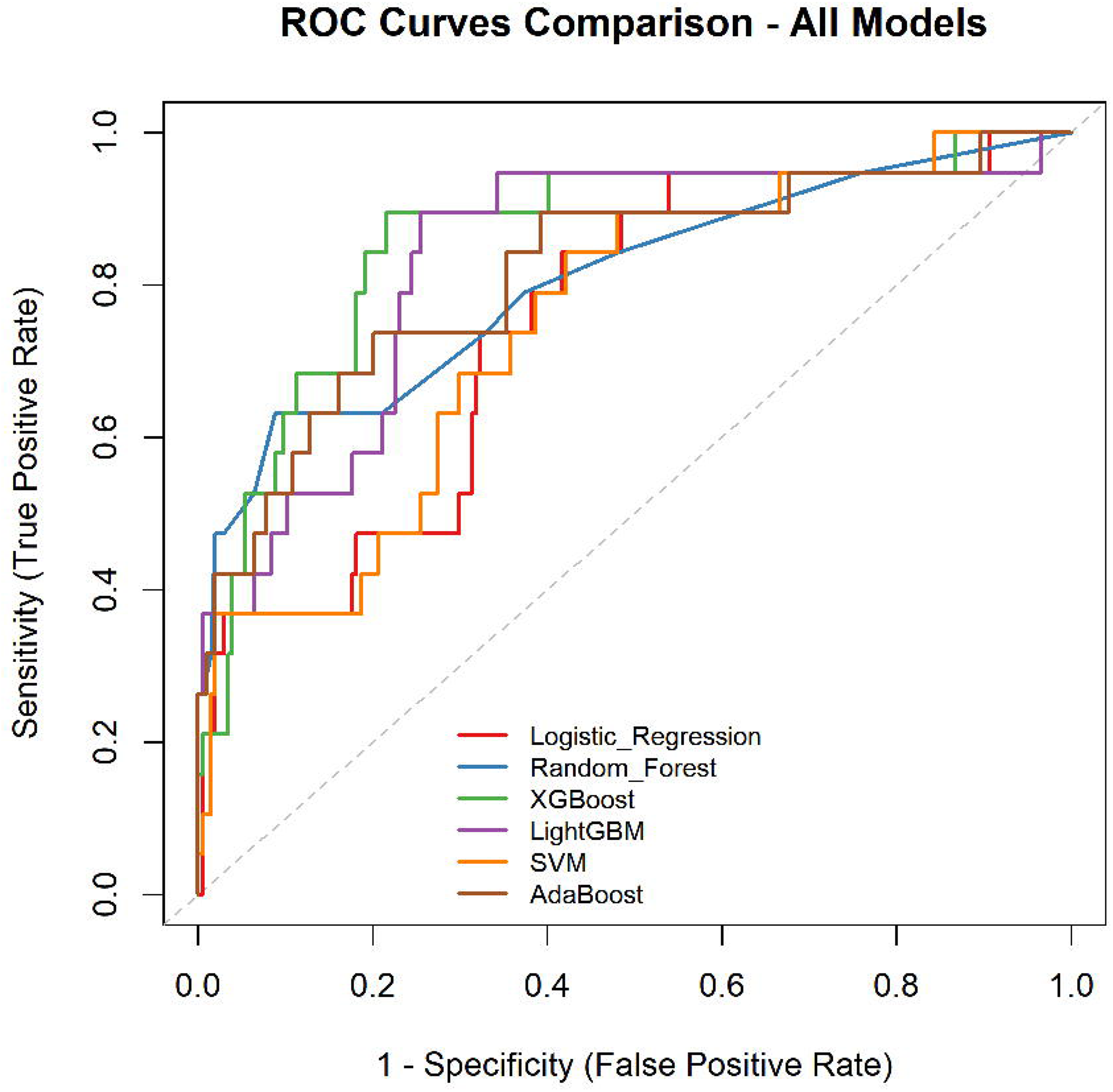
Comparison of ROC curves for each model. The plot illustrates the diagnostic ability of six different classifiers by plotting the True Positive Rate (Sensitivity) against the False Positive Rate (1 - Specificity). The diagonal dashed line represents the performance of a random classifier. The legend details the Area Under the Curve (AUC) values and 95% confidence intervals for each model: Logistic Regression (AUC = 0.75), Random Forest (AUC = 0.805), XGBoost (AUC = 0.863), LightGBM (AUC = 0.835), Support Vector Machine (SVM) (AUC = 0.751), and AdaBoost (AUC = 0.818). XGBoost demonstrated the highest discriminative power among the evaluated models.

**Table 7:**
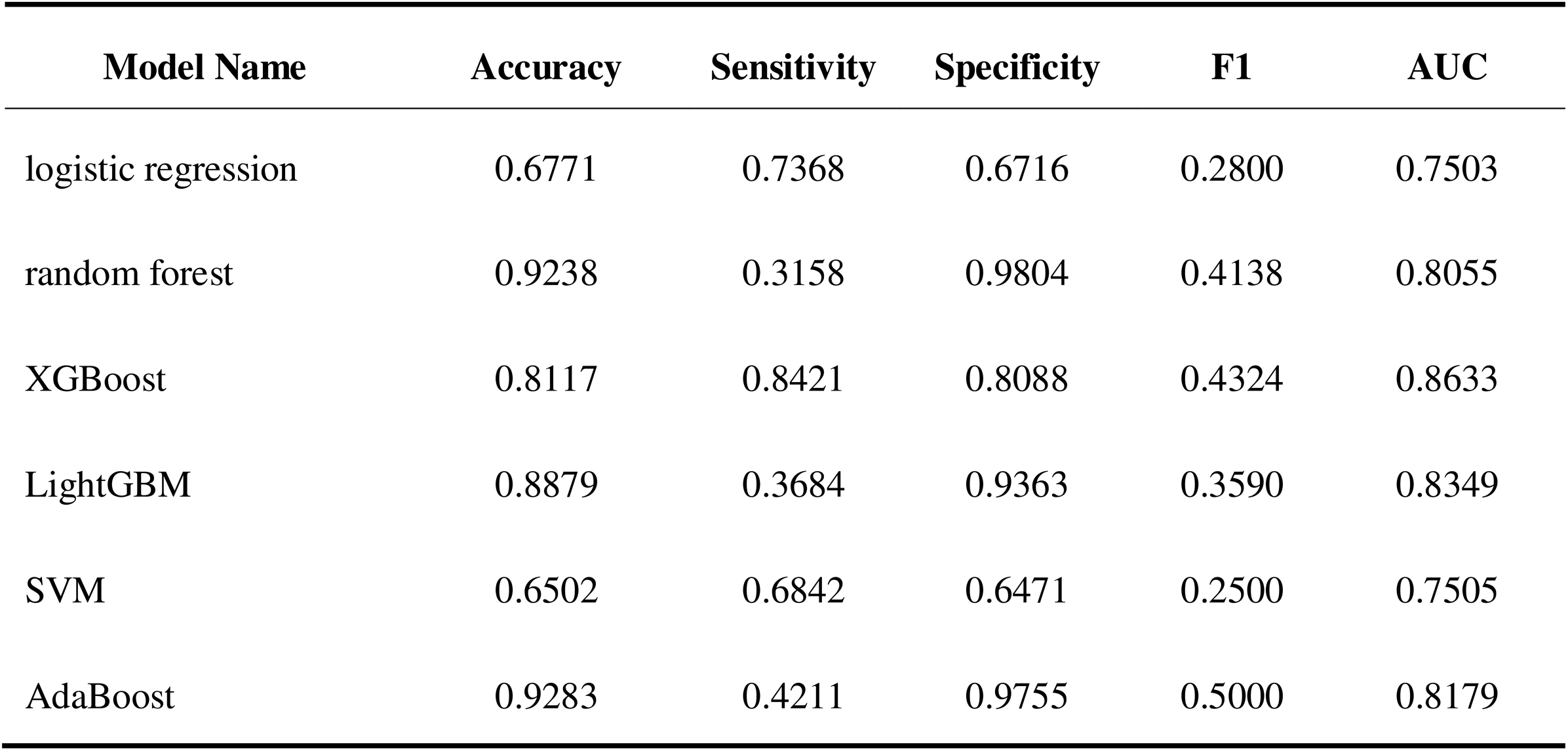
Performance Evaluation of Each Model’s Prediction Model.

##### 2. Explanatory analysis based on SHAP

Supplementary Figure 1 and figure 4 provide visual representations of the feature importance rankings derived from SHAP values in the XGBoost model. Specifically, Supplementary Figure 1 presents a swarm plot, while Figure 4 displays a feature importance plot. Both figures effectively illustrate the relative significance of each feature within the model. Age was identified as the most important predictor, followed by intraoperative blood loss, red blood cell suspension, BMI, GLU group, GAP group, reduction method group, MPV group, HDL-C group, ApoB group, kidney disease, and peripheral vascular disease(Figure 5).

**Figure 4:**
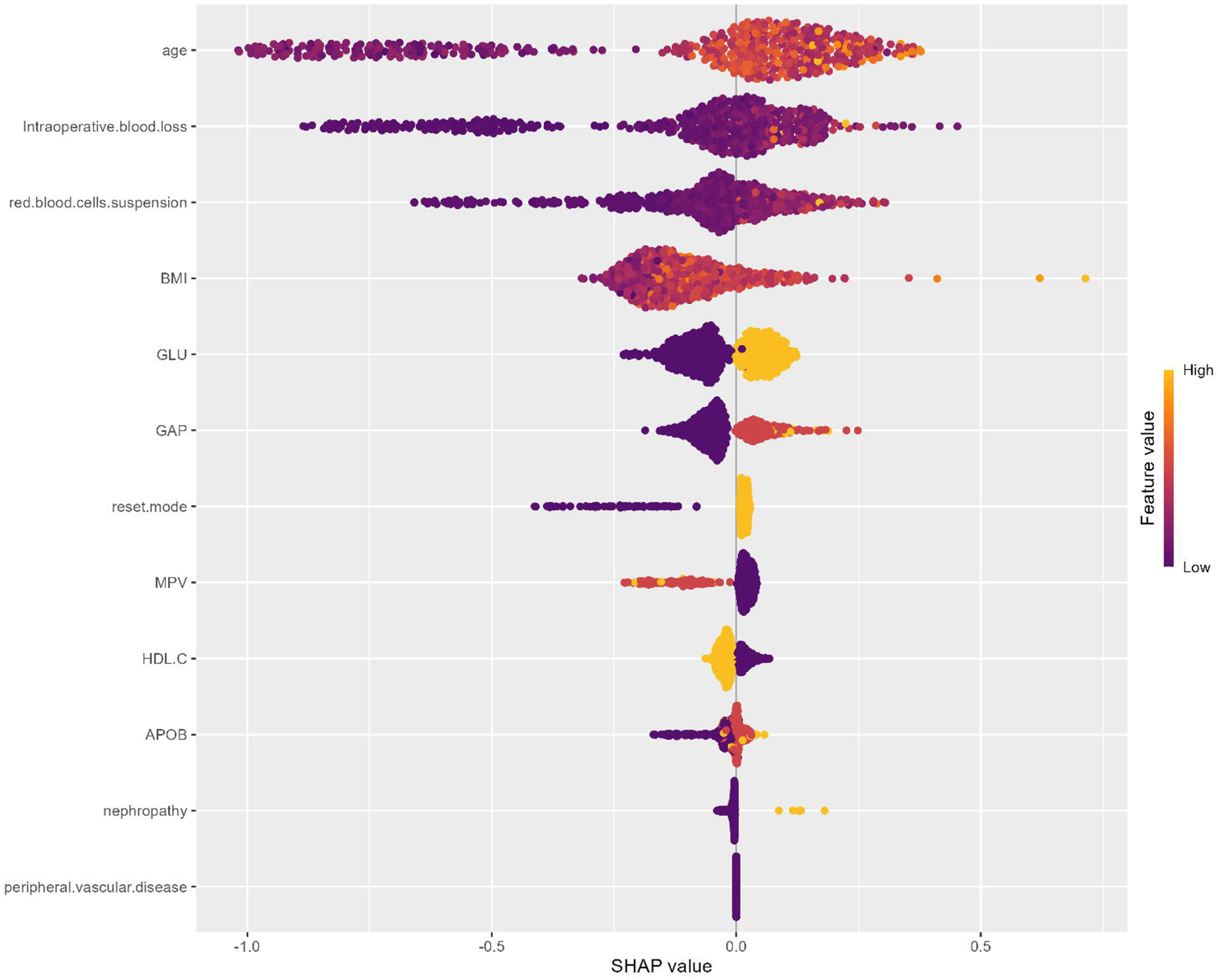
Summary plot of SHAP (SHapley Additive exPlanations) values, illustrating the impact of the top features on the model’s output

**Figure 5:**
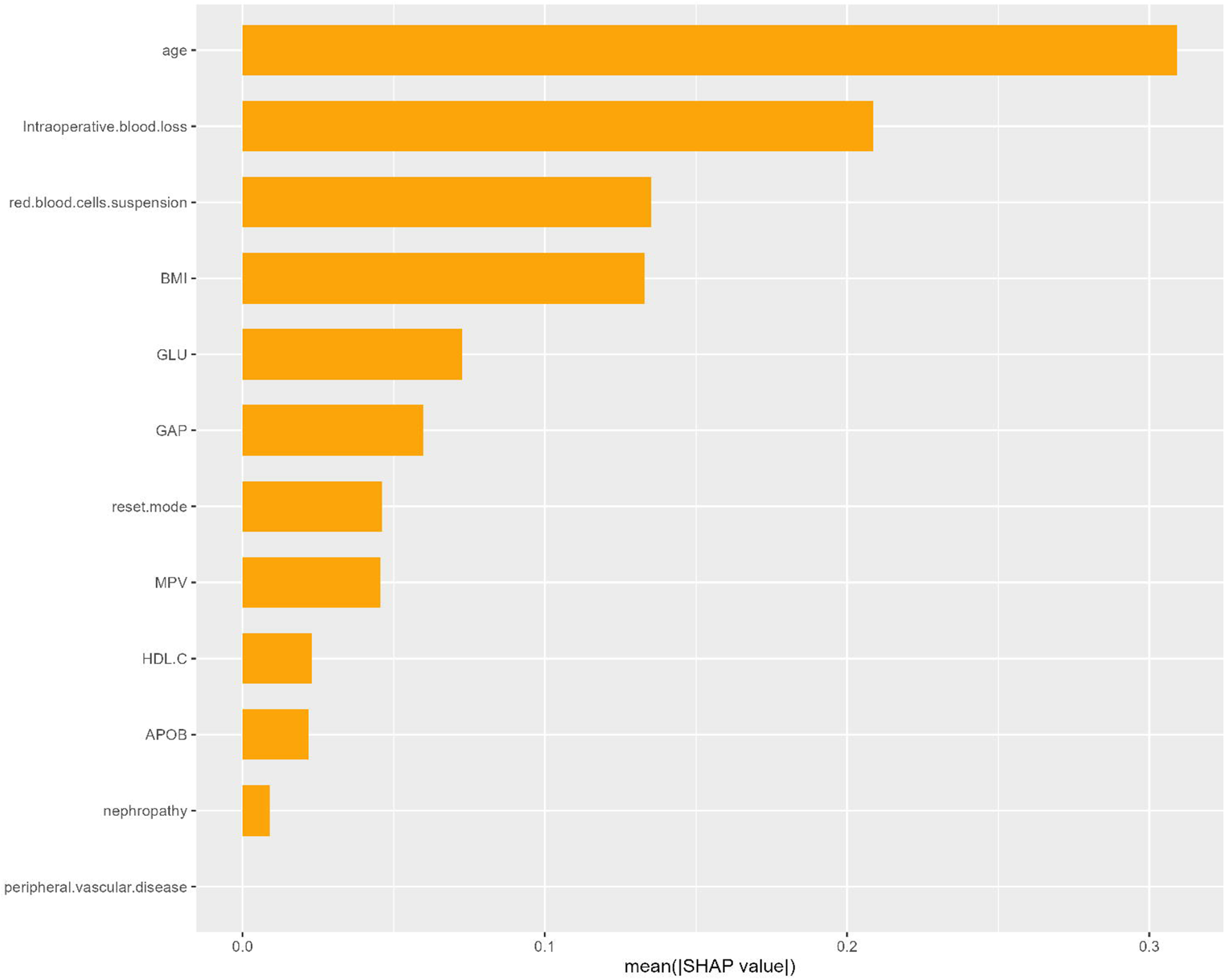
Feature Importance Ranking Chart Based on Shap Values

##### 3. Analysis based on SHAP waterfall chart and force-directed layout

Figure 6 demonstrates the utility of SHAP force plots for deconstructing individual predictions. These visualizations elucidate the contribution of each feature to a single sample’s outcome, where the color spectrum represents the direction of influence: features in yellow elevate the prediction score, while those in purple diminish it. The vertical axis enumerates the specific variables and their respective values for the given case. To illustrate this interpretability, force plots for representative positive and negative cases were generated. Analysis of a high-risk (positive) patient revealed that the model’s prediction was predominantly driven by several key factors. The patient’s age of 43 years was the most substantial contributor, increasing the prediction score by +0.269. This was closely followed by the administration of 2400 mL of packed red blood cells (+0.257) and an intraoperative blood loss of 700 mL (+0.16). Membership in GLU Group 1 provided a further, though smaller, positive influence (+0.103). Conversely, certain features moderated the overall risk. Membership in the GAP Group 0 exerted a slight protective effect (negative impact), partially offsetting the risk-enhancing factors. Other variables, such as HDL-C Group 0, MPV Group 0, reduction method 2, and ApoB Group 2, demonstrated minimal influence on the final prediction, despite their positive directional effect. The collective impact of the remaining features was negligible.

**Figure 6:**
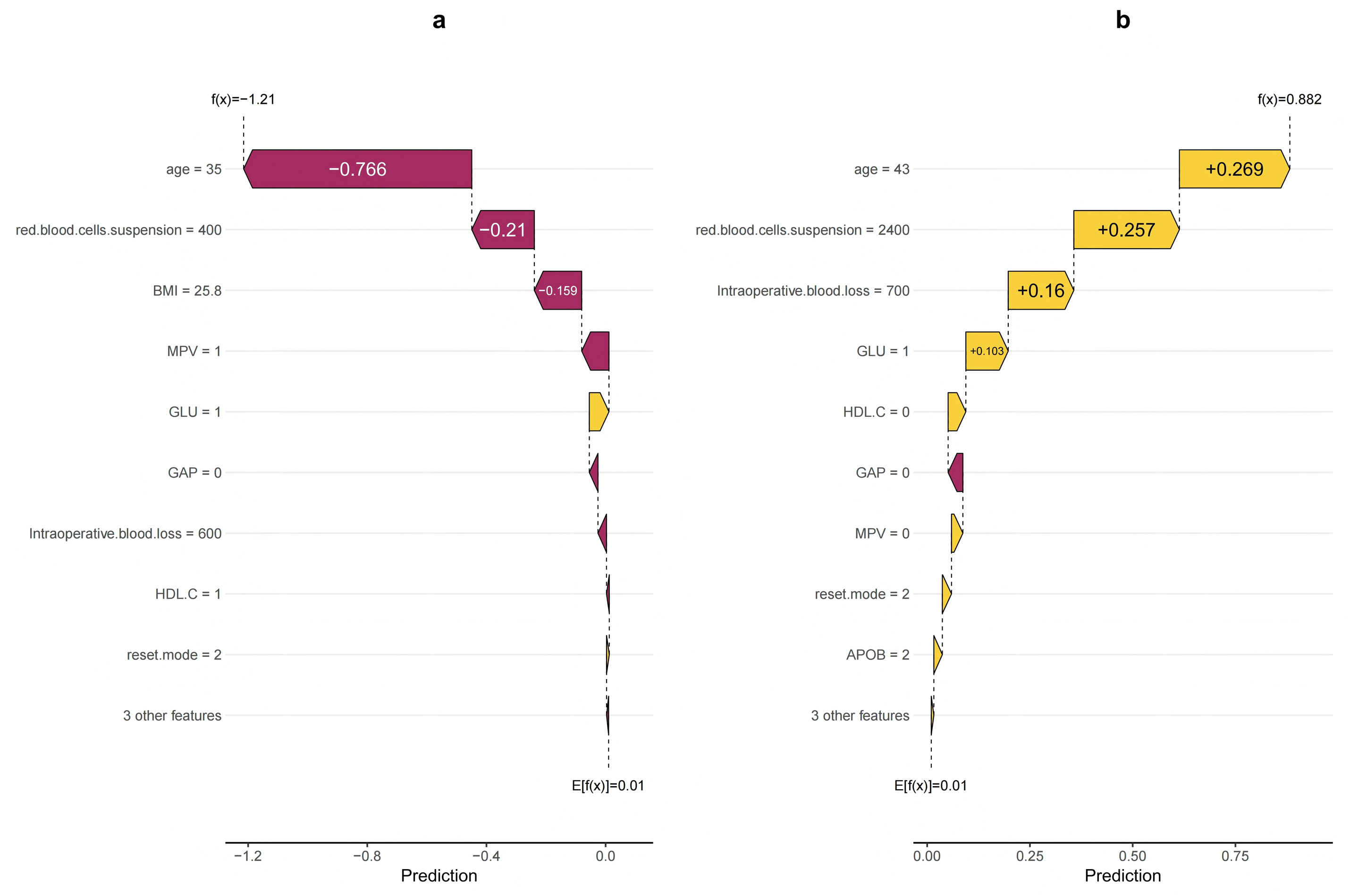
The Case’s Attemp.

**Figure 6a.** SHAP force plot for a negative case. This plot shows how individual features collectively push the prediction toward a low DVT risk. Younger age (35 years), lower transfusion volume (400 mL), and moderate BMI exert the strongest negative contributions, while only GLU Group 1 provides a small positive effect.

**Figure 6b.** SHAP force plot for a positive case. This plot illustrates the dominant factors driving a high predicted DVT risk. Older age (43 years), large-volume transfusion (2400 mL), and substantial intraoperative blood loss (700 mL) markedly increase the prediction score, whereas GAP Group 0 provides a minor protective influence.

The values associated with the features of negative sample patients are detailed as follows: Age 35 exerted the most significant negative impact on the prediction value, decreasing it by 0.766. The administration of 400 units of red blood cell suspension reduced the prediction value by 0.21, indicating a notable inhibitory effect. A BMI of 25.0 contributed a negative impact of 0.159, further decreasing the prediction value. The MPV group value of 1 reduced the prediction value by 0.0616, acting as a factor with a negative inhibitory effect.

Conversely, the GLU group value of 1, highlighted in yellow, positively influenced the prediction value, albeit with a relatively small amplitude. Features such as the GAP group value of 0 and intraoperative blood loss of 600, marked in purple, negatively affected the prediction value, though their influence was relatively minor, as indicated by the figure. Besides, features such as the HDL-C group value of 1 and the reduction method value of 2 yielded a discernible impact on the prediction value, with the degree of influence being relatively small, as illustrated in the figure. The remaining 3 had a negligible impact on the prediction value.

## Discussion

In recent years, the application of machine learning technology in the medical domain has notably expanded, particularly in thrombosis risk prediction, where it has demonstrated considerable benefits. Machine learning models can predict the likelihood of DVT after fractures or surgery, creating novel opportunities for tailored prevention. Empirical studies have shown that machine learning algorithms are proficient at handling substantial volumes of high-dimensional data, thereby facilitating the identification of novel risk factors and enhancing patient risk stratification^[11]^.

Machine learning models have demonstrated significant efficacy in predicting DVT across diverse surgical contexts. In tibial fracture surgery, Random Forest and SVM models achieved exceptional accuracy (AUC 0.99) and calibration ^[12]^. Similarly, ML approaches have proven superior to traditional risk scores like Caprini in laparoscopic surgery ^[13]^ and have emerged as effective non-invasive tools for early DVT detection in gastric cancer patients ^[14]^. The use of machine learning to predict DVT extends beyond the postoperative context. In patients experiencing spontaneous intracerebral hemorrhage, machine learning algorithms have been employed to forecast the risk of lower extremity DVT during hospitalization. The Light Gradient Boosting Machine (LGBM) model was found to exhibit a notable advantage in DVT prediction^[15]^. Furthermore, in oncology patients undergoing chemotherapy, machine learning models are utilized to evaluate the risk of catheter-related thrombosis. Despite the clinical application being constrained by complexity, the Bayesian learning model offers a simplified and robust alternative^[16]^.

As the utilization of machine learning algorithms in the medical field continues to expand, several researchers have endeavored to develop predictive models for DVT following fracture surgery using these advanced techniques. In the context of predicting DVT after various fractures or surgical procedures, numerous studies have demonstrated that machine learning models exhibit good performance in identifying significant risk factors and facilitating risk stratification. For instance, researchers have employed a range of algorithms for predicting DVT after knee and hip replacement surgeries, including XGBoost, RF, and SVM, integrating patient electronic health records (EHRs) to construct a highly accurate predictive model, achieving an AUC exceeding 0.92, which markedly surpasses the traditional Caprini score ^[17]^. Similarly, in the prediction of DVT following spinal surgery, the combination of Boruta and LASSO algorithms for feature selection enabled the random forest model to perform exceptionally well, accurately identifying patients at high risk ^[18]^. Furthermore, a study investigating DVT following surgical intervention for lower limb fractures compared various machine learning models, including XGBoost, Random Forest, and Logistic Regression. The results revealed that XGBoost exhibited superior performance with an AUC of 0.979, indicating robust predictive capability^[19]^.

Machine learning models offer substantial advantages over logistic regression in the context of large sample datasets, a perspective corroborated by numerous studies. Primarily, machine learning techniques excel at handling high-dimensional data and handling missing values. Contemporary machine learning approaches, including penalized regression, tree-based models, and deep learning, exhibit good performance in handling missing data within high-dimensional contexts, mitigating non-response bias, and demonstrating superiority in both simulation studies and practical applications^[20]^. Furthermore, in the analysis of large-scale neuroimaging datasets, machine learning effectively mitigates overfitting issues by integrating the predictive strengths of multiple classifiers, thereby enhancing the model’s generalization capabilities^[21]^.

Machine learning models consistently outperform traditional logistic regression in medical prediction, demonstrating superior discriminatory accuracy (higher AUC) for outcomes ranging from post-surgical mortality to disease severity ^[22,23]^. Their capacity to efficiently process large datasets and capture complex, non-linear variable interactions enhances model robustness and predictive reliability ^[24]^. Consequently, these algorithms improve clinical risk stratification by more accurately identifying high-risk patients and forecasting post-surgical complications^[25]^. In this study, we addressed a crucial clinical question: whether machine learning algorithms can improve the early prediction of postoperative non-occlusive deep vein thrombosis (PNO-DVT) in patients undergoing pelvic fracture surgery compared to traditional statistical methods. Although the traditional logistic regression model yielded a moderate AUC of 0.7503 (ranking fourth), the XGBoost model demonstrated superior discriminative performance. This finding aligns with a growing body of literature indicating that ensemble tree-based models consistently outperform logistic regression in complex clinical prediction tasks [22,23]. For instance, machine learning frameworks have achieved remarkable accuracy (with an AUC as high as 0.99) in tibial fracture cohorts [12] and have surpassed established clinical risk scores such as the Caprini model in laparoscopic surgical settings [13]. Although our model did not achieve near-perfect metrics in certain highly homogeneous fracture populations, its robust accuracy in the context of clinical heterogeneity and high risk associated with pelvic fractures underscores its practical relevance.

The superiority of XGBoost may stem from its ability to model nonlinear relationships and higher-order interactions among multifactorial predictive factors, such as intraoperative blood loss, BMI, blood glucose, and MPV, which are inherently ignored by linear regression. Clinically, these results suggest that integrating advanced ML algorithms into postoperative workflows can refine risk stratification, providing precise thrombosis prevention for high-risk patients while minimizing overtreatment for low-risk patients. Ultimately, this supports the shift in orthopedic trauma care from standardized DVT prevention protocols to personalized, data-driven clinical decision-making.

Additionally, in the field of deep vein thrombosis (DVT) risk prediction, the XGBoost model has shown better prediction accuracy and discriminatory ability than traditional tools in most studies. Taking the classic Caprini scoring system, which is widely used for clinical preoperative VTE risk stratification but relies on manually weighted clinical risk factors and fails to capture complex variable interactions, as an example, the XGBoost model, by automatically learning nonlinear relationships from large-scale clinical data, has demonstrated superior performance in multiple independent studies: a study on orthopedic inpatients showed its AUC reached 0.931, significantly higher than that of the Caprini model with improved accuracy and specificity, while another study on postoperative patients with digestive system tumors found it outperformed traditional statistical methods in feature selection and predictive performance. Moreover, XGBoost has good interpretability support (such as SHAP value analysis) to identify key predictive factors like D-dimer, fibrinogen and age for clinical individualized risk assessment, whereas traditional tools often struggle to dynamically integrate laboratory indicators with real-time patient condition changes.However, XGBoost still needs multi-center external validation to ensure generalization ability, while tools like Caprini are still widely used at the grassroots level due to their simple operation and no need for data platforms.

In patients with pelvic fractures, the severity of vascular injury is significantly correlated with prognosis. Research indicates that, even in cases of relatively isolated pelvic injuries, the severity of vascular injury is more strongly associated with clinical outcomes than the anatomical complexity of the fracture itself ^[26]^. Such vascular injuries not only elevate the risk of hemorrhage but may also initiate a series of inflammatory responses, thereby exacerbating the hypercoagulable state of the blood. In clinical practice, the management of trauma stress and inflammatory responses is crucial for improving the prognosis of patients with pelvic fractures. Research has shown that chronic psychological and social stress can affect the immune response through the β-adrenergic receptor signaling pathway, thereby influencing fracture healing^[26]^. Therefore, interventions targeting traumatic stress and inflammatory responses may alleviate hypercoagulability and improve overall patient prognosis. Accordingly, systemic inflammatory indicators SII, NLR, and PLR were incorporated into our analysis. Ultimately, none of these indicators were identified as significant predictors in this patient population.

Subsequently, SHAP values were employed to elucidate the probability of PNO-DVT as predicted by the XGBoost model. The analysis of SHAP values revealed that age was the most significant factor, followed by intraoperative blood loss, leukocyte removal, BMI, GLU group, GAP group, reduction method group, MPV group, HDL-C group, ApoB group, kidney disease, and peripheral vascular disease. Age and BMI, recognized as common cardiovascular risk factors, have been consistently associated with thrombosis in numerous studies. Current evidence indicates that older patients are at an increased risk for thrombotic events, potentially due to decreased vascular elasticity and the presence of atherosclerosis ^[28]^. Furthermore, an elevated BMI has been linked to metabolic syndrome and inflammatory responses, both of which may contribute to the development of thrombosis ^[29]^.

Intraoperative blood loss and the volume of suspended red blood cells transfused postoperatively are critical determinants influencing postoperative thrombosis. Significant intraoperative blood loss can result in hemodynamic instability, whereas excessive postoperative transfusion of suspended red blood cells may elevate blood viscosity, thereby augmenting the risk of thrombosis ^[30]^. These factors necessitate careful consideration in postoperative management to mitigate the risk of thrombosis.

Among laboratory test indicators, MPV is recognized as a marker of platelet activation and is closely associated with thrombosis. Empirical evidence suggests that patients with elevated MPV are more susceptible to venous thromboembolism (VTE), potentially due to the increased platelet volume, which enhances platelet reactivity and thus promotes thrombosis ^[31–32]^. Furthermore, integrating MPV with other hematological indicators can enhance the accuracy of thrombosis risk prediction ^[33]^.

Elevated glucose levels are significantly correlated with the incidence of DVT. Research indicates that hyperglycemia elevates the risk of thrombosis, potentially due to increased blood viscosity and vascular endothelial dysfunction associated with high glucose levels ^[34]^. Furthermore, individuals with diabetes are predisposed to thrombosis as a result of prolonged hyperglycemia, substantiating that glucose represents a risk factor for DVT ^[35]^.

Secondly, triglycerides and HDL-C are integral to lipid metabolism. Elevated triglyceride levels and reduced HDL-C levels have been linked to the incidence of DVT. Research indicates that dyslipidemia can damage the vascular endothelium and trigger inflammatory responses, thereby elevating the risk of thrombosis ^[36–37]^. Furthermore, low-density lipoprotein cholesterol (LDL-C) and the lymphocyte-to-lymphocyte ratio (LLR) have been documented as predictors of DVT, underscoring the significant role of lipid metabolism disorders in the pathogenesis of DVT ^[37]^.

Apolipoprotein B (APOB), the primary component of low-density lipoprotein, is significantly associated with atherosclerosis and thrombosis. Research indicates that elevated levels of APOB may elevate the risk of deep vein thrombosis (DVT) by promoting lipid deposition and vascular endothelial damage ^[38]^.

In conclusion, our machine learning model identified age, intraoperative blood loss, intraoperative and postoperative blood transfusion, body mass index (BMI), glucose (GLU), and mean platelet volume (MPV) as the most critical predictors of postoperative non-occlusive deep vein thrombosis (PNO-DVT) in patients with pelvic fractures across the entire dataset. The collection of these biomarkers facilitates the prediction of postoperative PNO-DVT risk, thereby informing clinical practice. Furthermore, we employed SHapley Additive exPlanations (SHAP) to enhance the model’s interpretability. SHAP values assist clinicians in providing personalized interventions for patients at high risk of DVT, allowing for tailored intervention strategies based on individual patient risk factors rather than a standardized treatment approach for all patients. This personalized approach promotes the efficient allocation of medical resources.

This study has several limitations that need to be considered. Firstly, the inherent retrospective design of this work introduces the possibility of selection bias and limits our ability to establish causality. From a methodological perspective, the performance of the developed machine learning model was not compared with existing clinical risk assessment tools such as Caprini or Autar scores, making it difficult to determine whether the proposed model offers significant advantages over standard clinical practice. Meanwhile, the model validation only employed a simple training-testing split method, without implementing external validation. This single validation strategy may lead to biases in the evaluation of model performance and fail to fully reflect the model’s stability and generalization ability under different data distributions. The lack of an external validation cohort also limits the generalizability of the research results, and the applicability of the model in other clinical settings or populations requires further verification. In addition, the simple data splitting method may not effectively assess the risk of overfitting of the model, and its reliability in real-world clinical practice still needs to be confirmed through larger-scale, multicenter prospective studies. Secondly, although we identified important predictive factors, residual confounding remains a distinct possibility; unmeasured perioperative variables such as specific activity protocols, variations in surgeon experience, or precise adherence to anticoagulation regimens were not captured in our dataset, and these variables may have independently influenced clinical outcomes. Furthermore, the diagnosis of DVT relies on clinical suspicion at the time and available imaging resources, increasing the potential for information bias. Lastly, regarding clinical application and generalizability, the current study results are based on a specific single-center dataset, and the practical value and workflow integration of the model remain unclear until it is validated in a prospective clinical setting. Therefore, the predictive model needs to be externally validated in different multicenter cohorts before it can be generalized to a broader patient population.

## Conclusion

In conclusion, this study demonstrates the utility of machine learning algorithms, particularly XGBoost, in predicting new-onset deep vein thrombosis (DVT) following pelvic fracture surgery. The analysis identified critical clinical predictors, including age, intraoperative blood loss, and body mass index, which consistently ranked high in importance across various models. The integration of these predictive tools into clinical practice offers a promising avenue for enhancing patient management. By enabling precise risk stratification, these models can assist clinicians in identifying high-risk patients early, facilitating timely preventative interventions, and ultimately improving post-operative outcomes.

## Data Availability

Data cannot be published without patients' consent. Researchers who are interested for academic need could contact the corresponding authors.

## Abbreviations

PNO-DVT: postoperative new-onset deep venous thrombosis
DVT: deep venous thrombosis
CV: continuous variable

## Declarations

### Ethics approval and consent to participate

According to the Helsinki Declaration, this study has been approved by the Ethics Committee of the Third Hospital of Hebei Medical University. Ethical batch number: KEO-2025-397-1. Before analysis, all data were anonymized, and since it was a retrospective study design, obtaining patient consent was not necessary.

## Data Availability Statement

Detailed data and R code for this article can be obtained from Professor Kuo Zhao.

## Consent for publication

All the authors have agreed to publish.

## Acknowledgements

Not applicable

## Clinical trial number

Not applicable

---

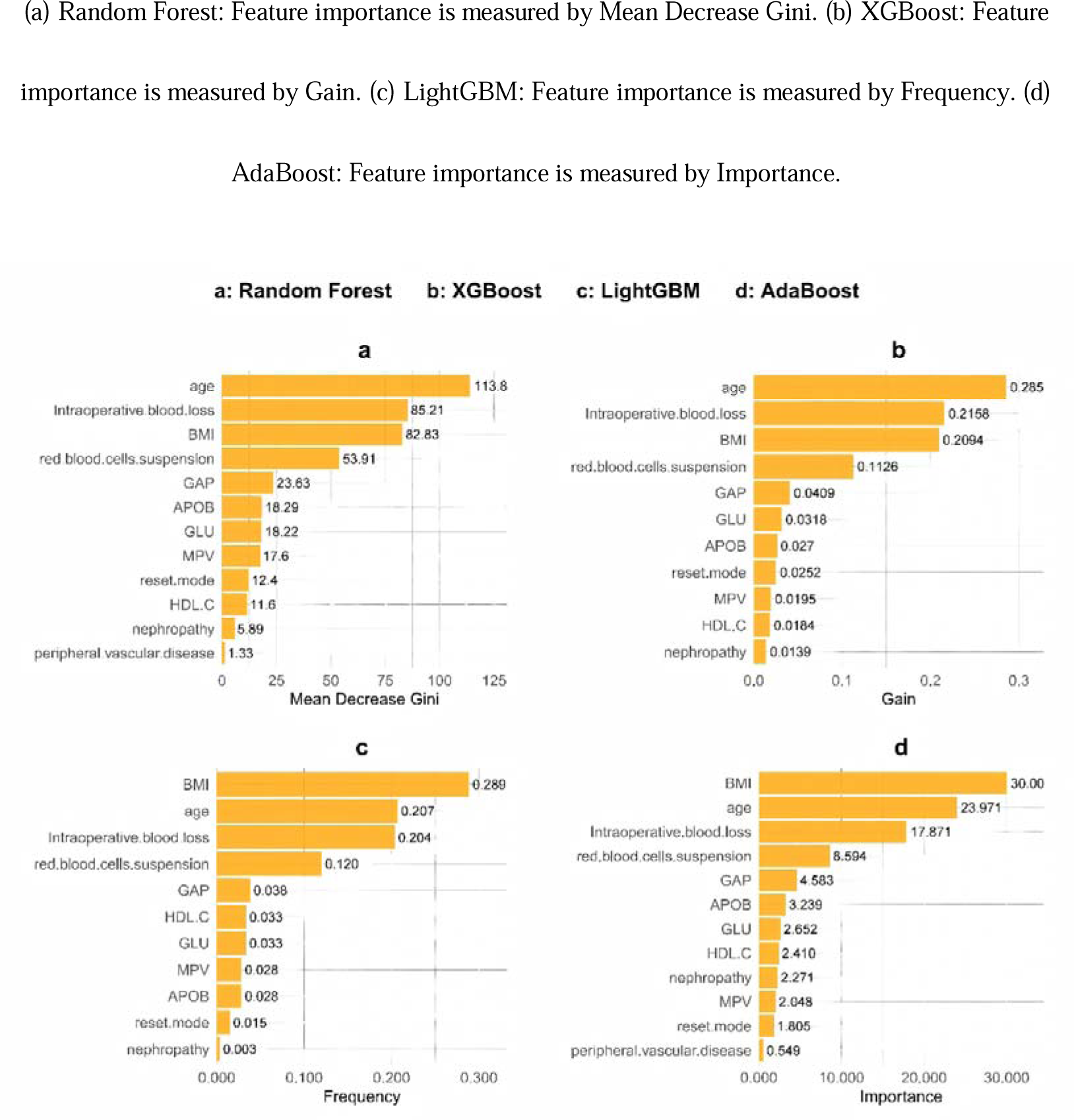

**Supplementary File S1.**
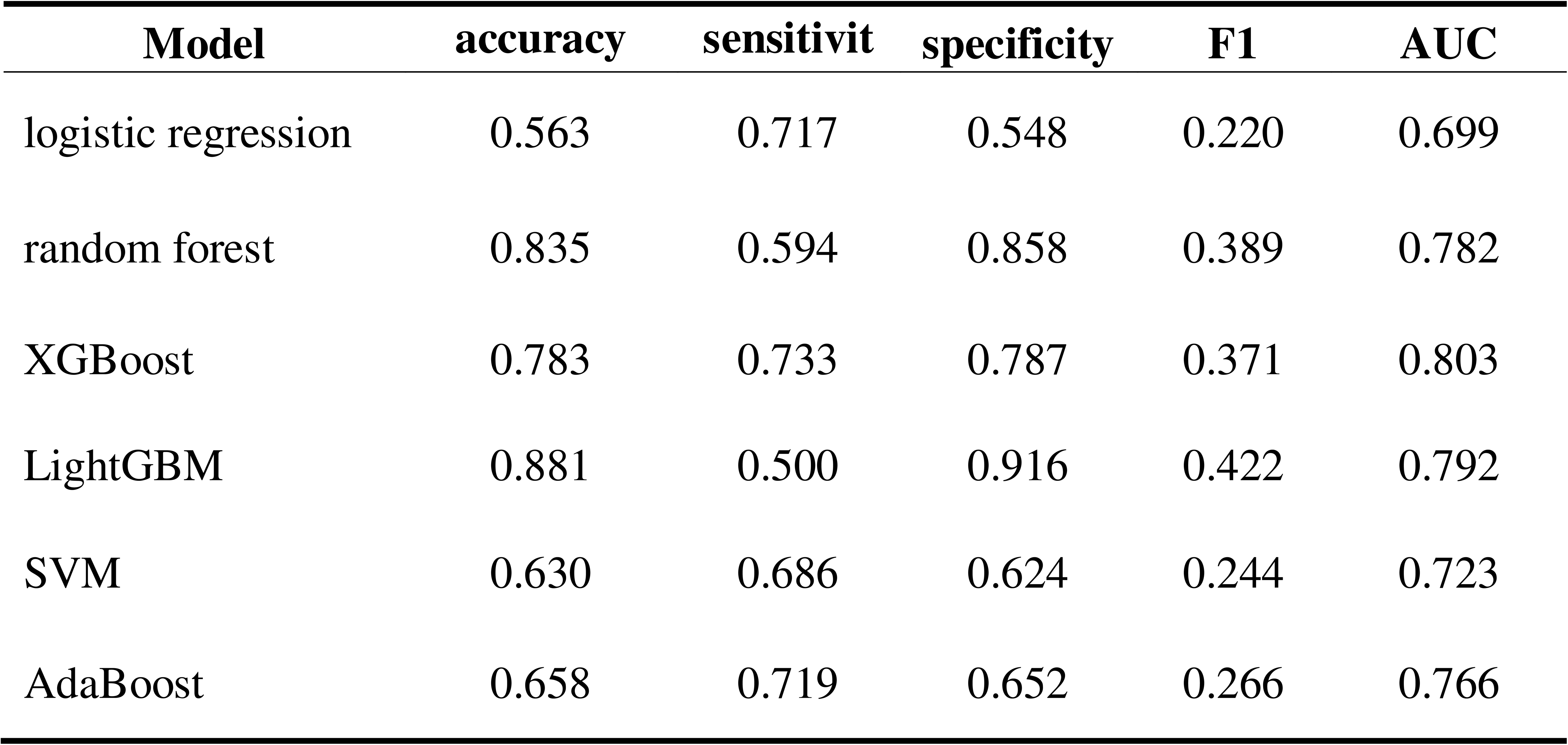
Average Performance Results of Each Model in 5-Fold Cross-Validation Samples.

## Supplementary File S1

To further assess the stability of the model, five-fold cross-validation was conducted on six algorithms, namely Logistic Regression, Random Forest, XGBoost, LightGBM, SVM, and AdaBoost. Each evaluation index was the average result of five cross-validations. As shown in the table below, the average AUC value of XGBoost over five times was 0.803, which remained the highest among all models. Moreover, it was higher than 0.8, indicating that this model has good discrimination ability and stable predictive performance. The model’s discriminatory efficacy is relatively reliable and has certain potential for clinical application.

## References

[1] . Chen H, Sun L, Kong X. Risk assessment scales to predict risk of lower extremity deep vein thrombosis among multiple trauma patients: a prospective cohort study. BMC Emerg Med. 2023 Dec 5;23(1):144. doi: 10.1186/s12873-023-00914-7. PMID: 38053029; PMCID: PMC10696745.

[2] . Chen Y, He J, Pan X. Prediction of risk factors for preoperative deep vein thrombosis in patients with pelvic fracture. Front Surg. 2025 Apr 28;12:1585460. doi: 10.3389/fsurg.2025.1585460. PMID: 40356946; PMCID: PMC12066781.

[3] . Wang H, Wu G, Chen CY, Qiu YY, Xie Y. Percutaneous screw fixation assisted by hollow pedicle finder for superior pubic ramus fractures. BMC Surg. 2022 Jun 3;22(1):216. doi: 10.1186/s12893-022-01659-z. PMID: 35658934; PMCID: PMC9166495.

[4] . Yang CS, Tan Z. Construction and validation of a predictive model for preoperative lower extremity deep vein thrombosis risk in elderly hip fracture patients: An observational study. Medicine (Baltimore). 2024 Sep 20;103(38):e39825. doi: 10.1097/MD.0000000000039825. PMID: 39312315; PMCID: PMC11419451.

[5] . Orak F, Saadat M, Saki Malehi A, Behdarvandan A, Esfandiarpour F. Comparison of the Pauda and the Autar DVT Risk Assessment Scales in Prediction of Venous Thromboembolism in ICU Patients. Med J Islam Repub Iran. 2024 Apr 30;38:48. doi: 10.47176/mjiri.38.48. PMID: 39399622; PMCID: PMC11469705.

[6] . Shekarchian S, Notten P, Barbati ME, Van Laanen J, Piao L, Nieman F, Razavi MK, Lao M, Mees B, Jalaie H. Development of a prediction model for deep vein thrombosis in a retrospective cohort of patients with suspected deep vein thrombosis in primary care. J Vasc Surg Venous Lymphat Disord. 2022 Sep;10(5):1028–1036.e3. doi: 10.1016/j.jvsv.2022.04.009. Epub 2022 May 27. PMID: 35644336.

[7] . Tang Z, Li N, Tian Y. A nomogram for predicting risk factors for lower limb deep venous thrombosis in elderly postoperative patients with severe traumatic brain injury in the intensive care unit. Phlebology. 2025 Jul;40(6):446–455. doi: 10.1177/02683555251332988. Epub 2025 Apr 10. PMID: 40205921.

[8] . Wu L, Zhao Y, Yao G, Li X, Zhao X. Prediction and Analysis of Risk Factors for Lower Extremity Deep Vein Thrombosis After Craniotomy in Patients with Primary Brain Tumors: A Machine Learning Approach. Turk Neurosurg. 2025;35(4):636–643. doi: 10.5137/1019-5149.JTN.47938-24.3. PMID: 40577511.

[9] . Qiu W, Cui P, Li S, Tang Z, Chen J, Wang J, Li Y. Machine learning models predict risk of lower extremity deep vein thrombosis in hospitalized patients with spontaneous intracerebral hemorrhage. Sci Rep. 2025 Jul 10;15(1):24932. doi: 10.1038/s41598-025-10905-2. PMID: 40640445; PMCID: PMC12246041.

[10] . Li R, Chen S, Xia J, Zhou H, Shen Q, Li Q, Dong Q. Predictive modeling of deep vein thrombosis risk in hospitalized patients: A Q-learning enhanced feature selection model. Comput Biol Med. 2024 Jun;175:108447. doi: 10.1016/j.compbiomed.2024.108447. Epub 2024 Apr 12. PMID: 38691912.

[11] . Chrysafi P, Lam B, Carton S, Patell R. From Code to Clots: Applying Machine Learning to Clinical Aspects of Venous Thromboembolism Prevention, Diagnosis, and Management. Hamostaseologie. 2024 Dec;44(6):429–445. doi: 10.1055/a-2415-8408. Epub 2024 Dec 10. PMID: 39657652.

[12] . Baki H, Özçelik İB. Machine Learning-Based Prediction of Postoperative Deep Vein Thrombosis Following Tibial Fracture Surgery. Diagnostics (Basel). 2025 Jul 16;15(14):1787. doi: 10.3390/diagnostics15141787. PMID: 40722536; PMCID: PMC12293441.

[13] . Yang SZ, Peng MH, Lin Q, Guan SW, Zhang KL, Yu HB. A machine learning-based predictive model for the occurrence of lower extremity deep vein thrombosis after laparoscopic surgery in abdominal surgery. Front Surg. 2025 May 30;12:1502944. doi: 10.3389/fsurg.2025.1502944. PMID: 40520687; PMCID: PMC12162540.

[14] . Zeng Y, Chen Y, Zhu D, Xu J, Zhang X, Ying H, Song X, Zhou R, Wang Y, Yu F. Machine learning assisted radiomics in predicting postoperative occurrence of deep venous thrombosis in patients with gastric cancer. BMC Cancer. 2025 Feb 7;25(1):220. doi: 10.1186/s12885-025-13630-1. PMID: 39920636; PMCID: PMC11806839.

[15] . Qiu W, Cui P, Li S, Tang Z, Chen J, Wang J, Li Y. Machine learning models predict risk of lower extremity deep vein thrombosis in hospitalized patients with spontaneous intracerebral hemorrhage. Sci Rep. 2025 Jul 10;15(1):24932. doi: 10.1038/s41598-025-10905-2. PMID: 40640445; PMCID: PMC12246041.

[16] . An T, Han H, Xie J, Wang Y, Zhao Y, Jia H, Wang Y. Enhancing prediction and stratifying risk: machine learning and bayesian-learning models for catheter-related thrombosis in chemotherapy patients. BMC Cancer. 2025 Mar 27;25(1):552. doi: 10.1186/s12885-025-13946-y. PMID: 40148861; PMCID: PMC11948715.

[17] . Wang X, Xi H, Geng X, Li Y, Zhao M, Li F, Li Z, Ji H, Tian H. Artificial Intelligence-Based Prediction of Lower Extremity Deep Vein Thrombosis Risk After Knee/Hip Arthroplasty. Clin Appl Thromb Hemost. 2023 Jan-Dec;29:10760296221139263. doi: 10.1177/10760296221139263. PMID: 36596268; PMCID: PMC9830569.

[18] . Wu X, Wang Z, Zheng L, Yang Y, Shi W, Wang J, Liu D, Zhang Y. Construction and verification of a machine learning-based prediction model of deep vein thrombosis formation after spinal surgery. Int J Med Inform. 2024 Dec;192:105609. doi: 10.1016/j.ijmedinf.2024.105609. Epub 2024 Aug 30. PMID: 39260049.

[19] . Wei C, Wang J, Yu P, Li A, Xiong Z, Yuan Z, Yu L, Luo J. Comparison of different machine learning classification models for predicting deep vein thrombosis in lower extremity fractures. Sci Rep. 2024 Mar 22;14(1):6901. doi: 10.1038/s41598-024-57711-w. PMID: 38519523; PMCID: PMC10960026.

[20] . Chen S, Xu C. Handling high-dimensional data with missing values by modern machine learning techniques. J Appl Stat. 2022 May 1;50(3):786–804. doi: 10.1080/02664763.2022.2068514. PMID: 36819079; PMCID: PMC9930810.

[21] . Lanka P, Rangaprakash D, Dretsch MN, Katz JS, Denney TS Jr, Deshpande G. Supervised machine learning for diagnostic classification from large-scale neuroimaging datasets. Brain Imaging Behav. 2020 Dec;14(6):2378–2416. doi: 10.1007/s11682-019-00191-8. PMID: 31691160; PMCID: PMC7198352.

[22] . Leonard G, South C, Balentine C, Porembka M, Mansour J, Wang S, Yopp A, Polanco P, Zeh H, Augustine M. Machine Learning Improves Prediction Over Logistic Regression on Resected Colon Cancer Patients. J Surg Res. 2022 Jul;275:181–193. doi: 10.1016/j.jss.2022.01.012. Epub 2022 Mar 11. PMID: 35287027.

[23] . Ye J, Hua M, Zhu F. Machine Learning Algorithms are Superior to Conventional Regression Models in Predicting Risk Stratification of COVID-19 Patients. Risk Manag Healthc Policy. 2021 Jul 29;14:3159–3166. doi: 10.2147/RMHP.S318265. PMID: 34349576; PMCID: PMC8328384.

[24] . Bailly A, Blanc C, Francis É, Guillotin T, Jamal F, Wakim B, Roy P. Effects of dataset size and interactions on the prediction performance of logistic regression and deep learning models. Comput Methods Programs Biomed. 2022 Jan;213:106504. doi: 10.1016/j.cmpb.2021.106504. Epub 2021 Oct 28. PMID: 34798408.

[25] . Michelsen C, Jørgensen CC, Heltberg M, Jensen MH, Lucchetti A, Petersen PB, Petersen T, Kehlet H; Center for Fast-track Hip Knee Replacement Collaborative group; Madsen F, Hansen TB, Gromov K, Jakobsen T, Varnum C, Overgaard S, Rathsach M, Hansen L. Machine-learning vs. logistic regression for preoperative prediction of medical morbidity after fast-track hip and knee arthroplasty-a comparative study. BMC Anesthesiol. 2023 Nov 29;23(1):391. doi: 10.1186/s12871-023-02354-z. PMID: 38030979; PMCID: PMC10685559.

[26] . Wu YT, Cheng CT, Tee YS, Fu CY, Liao CH, Hsieh CH. Pelvic injury prognosis is more closely related to vascular injury severity than anatomical fracture complexity: the WSES classification for pelvic trauma makes sense. World J Emerg Surg. 2020 Aug 17;15(1):48. doi: 10.1186/s13017-020-00328-x. PMID: 32807185; PMCID: PMC7433075.

[27] . Haffner-Luntzer M, Foertsch S, Fischer V, Prystaz K, Tschaffon M, Mödinger Y, Bahney CS, Marcucio RS, Miclau T, Ignatius A, Reber SO. Chronic psychosocial stress compromises the immune response and endochondral ossification during bone fracture healing via β-AR signaling. Proc Natl Acad Sci U S A. 2019 Apr 23;116(17):8615–8622. doi: 10.1073/pnas.1819218116. Epub 2019 Apr 4. PMID: 30948630; PMCID: PMC6486758.

[28] . Rong X, Jiang L, Qu M, Yang S, Wang K, Jiang L. Risk factors and characteristics of ischemic stroke in patients with immune thrombocytopenia: A retrospective cohort study. J Stroke Cerebrovasc Dis. 2022 Oct;31(10):106693. doi: 10.1016/j.jstrokecerebrovasdis.2022.106693. Epub 2022 Aug 31. PMID: 36054971.

[29] . Hansen ES, Edvardsen MS, Aukrust P, Ueland T, Hansen JB, Brækkan SK, Morelli VM. Combined effect of high factor VIII levels and high mean platelet volume on the risk of future incident venous thromboembolism. J Thromb Haemost. 2023 Oct;21(10):2844–2853. doi: 10.1016/j.jtha.2023.06.022. Epub 2023 Jun 29. PMID: 37393000.

[30] . Zhang YM, Chen W, Wei HL, Tang XH, Xie FH, Wang RX. Analysis of predictive factors of thrombosis in autogenous arteriovenous fistula. J Vasc Access. 2024 Jul;25(4):1134–1139. doi: 10.1177/11297298221151135. Epub 2023 Jan 27. PMID: 36707987.

[31] . Edvardsen MS, Hansen ES, Hindberg K, Morelli VM, Ueland T, Aukrust P, Brækkan SK, Evensen LH, Hansen JB. Combined effects of plasma von Willebrand factor and platelet measures on the risk of incident venous thromboembolism. Blood. 2021 Dec 2;138(22):2269–2277. doi: 10.1182/blood.2021011494. PMID: 34161566.

[32] . Wang Z, Chen X, Wu J, Zhou Q, Liu H, Wu Y, Liu S, Liu Y. Low Mean Platelet Volume is Associated with Deep Vein Thrombosis in Older Patients with Hip Fracture. Clin Appl Thromb Hemost. 2022 Jan-Dec;28:10760296221078837. doi: 10.1177/10760296221078837. PMID: 35157546; PMCID: PMC8848069.

[33] . Jakobsen L, Frischmuth T, Brækkan SK, Hansen JB, Morelli VM. Joint Effect of Multiple Prothrombotic Genotypes and Mean Platelet Volume on the Risk of Incident Venous Thromboembolism. Thromb Haemost. 2022 Nov;122(11):1911–1920. doi: 10.1055/a-1863-2052. Epub 2022 May 26. PMID: 35617954.

[34] . Liu X, Li T, Xu H, Wang C, Ma X, Huang H, Hu Y, Chu H. Hyperglycemia may increase deep vein thrombosis in trauma patients with lower limb fracture. Front Cardiovasc Med. 2022 Sep 8;9:944506. doi: 10.3389/fcvm.2022.944506. PMID: 36158801; PMCID: PMC9498976.

[35] . Hang L, Haibier A, Kayierhan A, Abudurexiti T. Risk factors for deep vein thrombosis of the lower extremity after total hip arthroplasty. BMC Surg. 2024 Sep 11;24(1):256. doi: 10.1186/s12893-024-02561-6. PMID: 39261801; PMCID: PMC11389418.

[36] . Abdelmalik BHA, Leslom MMA, Gameraddin M, Alshammari QT, Hussien R, Alyami MH, Salih M, Yousef M, Yousif E. Assessment of Lower Limb Deep Vein Thrombosis: Characterization and Associated Risk Factors Using Triplex Doppler Imaging. Vasc Health Risk Manag. 2023 May 4;19:279–287. doi: 10.2147/VHRM.S409253. PMID: 37168880; PMCID: PMC10166097.

[37] . Guo H, Li C, Wu H, Ma M, Zhu R, Wang M, Yang B, Pan N, Zhu Y, Wang J. Low-density lipoprotein cholesterol-to-lymphocyte count ratio (LLR) is a promising novel predictor of postoperative new-onset deep vein thrombosis following open wedge high tibial osteotomy: a propensity score-matched analysis. Thromb J. 2024 Jul 16;22(1):64. doi: 10.1186/s12959-024-00635-2. PMID: 39014396; PMCID: PMC11250942.

[38] . Gu H, Yang F, Xie H, Li M, Wang Z, Sheng L. Serum VEGF, P-Selectin, HDL-C, Platelet Index, and Coagulation Function Index in Deep Vein Thrombosis after Traumatic Fracture. Clin Lab. 2024 Feb 1;70(2). doi: 10.7754/Clin.Lab.2023.230425. PMID: 38345981.

